# Development and Validation of the SHERLOC Model: A Disease Model for Non-Small Cell Lung Cancer

**DOI:** 10.64898/2026.11.27.26369000

**Authors:** David C. Griffin, Syed Irteza Abbas Shamsi, Amanda AW. Gibson, Michelle L. Dean, Vishal Navani, Winson Y. Cheung, Alain Tremblay, Eldon Spackman

## Abstract

**Objective:** Lung Cancer is the leading cause of cancer-related death internationally, with Non-Small Cell Lung Cancer (NSCLC) representing more than 85% of diagnoses. Despite the large number of economic evaluations in NSCLC, most utilize decision-problem-specific models. To address this, we developed and validated a microsimulation model for NSCLC and demonstrated it by performing a case study assessing the FLAURA2 and MARIPOSA trials.

**Methods:** Utilizing survival data from a cohort derived from the Glans-Look Lung Cancer Research (GLR) database, publicly available list prices, and relevant literature, a microsimulation with a 25-year time horizon was developed in R that estimates average costs, quality-adjusted life years (QALYs), and life-years (LYs). The model was validated internally and externally through calculated mean absolute percent error (MAPE), mean absolute error (MAE), and R^2^. Case study results were compared to Canada’s Drug Agency’s (CDA-AMC) pharmacoeconomic report.

**Results:** NSCLC was estimated to cost $205,470 (95% CI: $200,950-$209,990) with average QALYs per case of 3.39 (95% CI: 3.31-3.46). Internal validation had an R^2^ of 0.9721 against the GLR cohort and an R^2^ of 0.8795 externally, with an MAPE of 13.9%. Incremental costs for osimertinib plus platinum pemetrexed, and amivantamab plus lazertinib against osimertinib were $110,525 (95% CI: $106,969-$114,082) and $186,855 (95% CI: $181,060-192,651), and incremental QALYs were 0.413 (95% CI: 0.389-0.436) and 0.483 (95% CI: 0.461-0.504), slightly lower than the CDA-AMCs deterministic/probabilistic findings. Incremental cost-effectiveness ratios were $267,908 and $387,223.

**Conclusions:** The model represents a transparent, internally and externally validated foundation for the assessment of preventative and oncology health technology interventions in lung cancer.

## Introduction

Lung cancer is the most common cause of cancer-related mortality in the world.^1^ Of these cancers, Non-Small-Cell Lung Cancer (NSCLC) makes up a growing 85%+ of cases.^1^ This is due to the shifting causes of lung cancer. Currently, while tobacco is attributable for a majority of cases, the proportion attributable to non-tobacco-related causes (radon, PM_2.5_, and other occupational carcinogens) represents a growing proportion of lung cancers.^2^ This proportion is growing, with age-standardized incidence rates for small cell lung cancer showing an ongoing downward trend while adenocarcinoma shows a stable, if not increasing, trend.^3^ Advancements with novel immunotherapies, such as pembrolizumab and tyrosine kinase inhibitors, such as osimertinib and alectinib, have become the standard of care in modern healthcare systems.^4–7^ As would be expected, survival rates have consistently shown an upward trend in line with advances in treatment. Similarly, as survival rates improve, the economic burden of lung cancer would be expected to grow through extended survival and higher prices for therapies.^8,9^ As new novel therapies and health technologies are developed, the importance of validated models capable of assessing novel therapies and preventive interventions is clear.

The majority of economic evaluations utilize decision-problem-specific models that use published data to evaluate interventions with only one use case specific to the intervention of interest.^9,10^ With sometimes multiple potential therapies for one decision problem, this would be assessed separately with models that can have differing structures, assumptions, and evidence, making informed judgements sometimes difficult for health technology assessment bodies.^8,11^ Further, the majority of published models in NSCLC cost-effectiveness analyses have been found to utilize simple 3-state models or partition state models, and only a minority report internal or external validity.^9^ The lack of validation or reporting of validation raises questions about the accuracy and validity of models being used for economic evaluations in NSCLC.^9^ However, the use of probabilistic sensitivity analyses was generally widespread, as is recommended for health technology assessments.^9,12^

Resultingly, decision makers are faced with the challenge of making recommendations for therapies based on evidence with differing assumptions and models addressing a singular decision problem. The piecewise nature and limited scope of these models further means that the effect of multiple interventions cannot be assessed simultaneously. Models that can assess interventions in the full course of the disease have been proposed as a solution to these shortcomings.^10^ This requires a model capable of assessing technologies aimed at prevention and emerging novel oncology therapies.

Internationally, models for NSCLC tend not to meet the above requirements while also meeting recommended standards and being open-source.^12^ Currently, an example of a detailed NSCLC model is the OncoSim-Lung model, which has been validated against the NLST trial and has received extensive work over time, although publicly available information indicates some parameters are currently outdated.^13–15^ However, the authors have noted that a limitation of the model is that it cannot perform probabilistic sensitivity analyses through second-order simulations, as is recommended by health technology assessment bodies in economic evaluations to fully assess parameter uncertainty in decision problems.^16^ Further, while OncoSim-Lung has been praised as a tool for policy recommendations in lung cancer screening, limitations were noted in the models’ coding transparency and assessment of non-preventative technologies.^17^ The complexity of models such as OncoSim-Lung means that they become akin to “black boxes”. A lack of transparency in code-opaque models can impair the credibility and reproducibility of results, along with hindering critical evaluation steps.^18–20^

To address these, we developed a discrete-time individual state transition model, also known as a microsimulation, capable of assessing interventions at multiple stages of the disease process, named the “Simulation for the Health Economics of Routine NSCLC Oncology in Canada” model or SHERLOC model. This model was informed by real-world survival data, current standards of care and up-to-date costs. We aimed to provide a consistent open-source model for the assessment of preventative and oncology health technology interventions in NSCLC, initially designed for the Canadian setting. To demonstrate the NSCLC models’ usefulness in assessing multiple interventions for one decision problem and to further validate the model, we performed a comparative cost-effectiveness assessment from a healthcare payer perspective of the recently published FLAURA2 and MARIPOSA trials.^21,22^ These trials compare osimertinib plus chemotherapy to osimertinib monotherapy and amivantimab plus lazertinib in the first-line *EGFR*-Mutated Advanced NSCLC setting, and the evaluation results can be compared to the evaluation made by Canada’s Drug Agency (CDA-AMC).^8,21,22^

## Methods

### Overview

The SHERLOC model was developed with the goal of producing a model capable of assessing interventions throughout the disease pathway (Figure 1), designed to best model how NSCLC is currently treated. The model is informed by real-world survival data, published health utilities, and best practice standards of care to model resource usage. The model defaults to a 25-year time horizon with a 1-month cycle to capture the full long-term effects of interventions, but can be varied anywhere from the full 25 years to 1 month. The model outputs are costs in 2025 $CAD from a public payer perspective, Quality-Adjusted Life Years (QALYs) and Life Years (LYs) with a standard discount rate of 1.5% applied.^12,23^ The model and analysis were developed in line with CADTH (now CDA-AMC) guidelines and reported in accordance with the CHEERS (Consolidated Health Economic Evaluation Reporting Standards 2022) guidelines.^12,24^ Chart review was approved by the Health Research Ethics Board of Alberta, Cancer Committee (HREBA.CC-16-0574), in accordance with regional guidelines and regulations.

**Figure 1:**
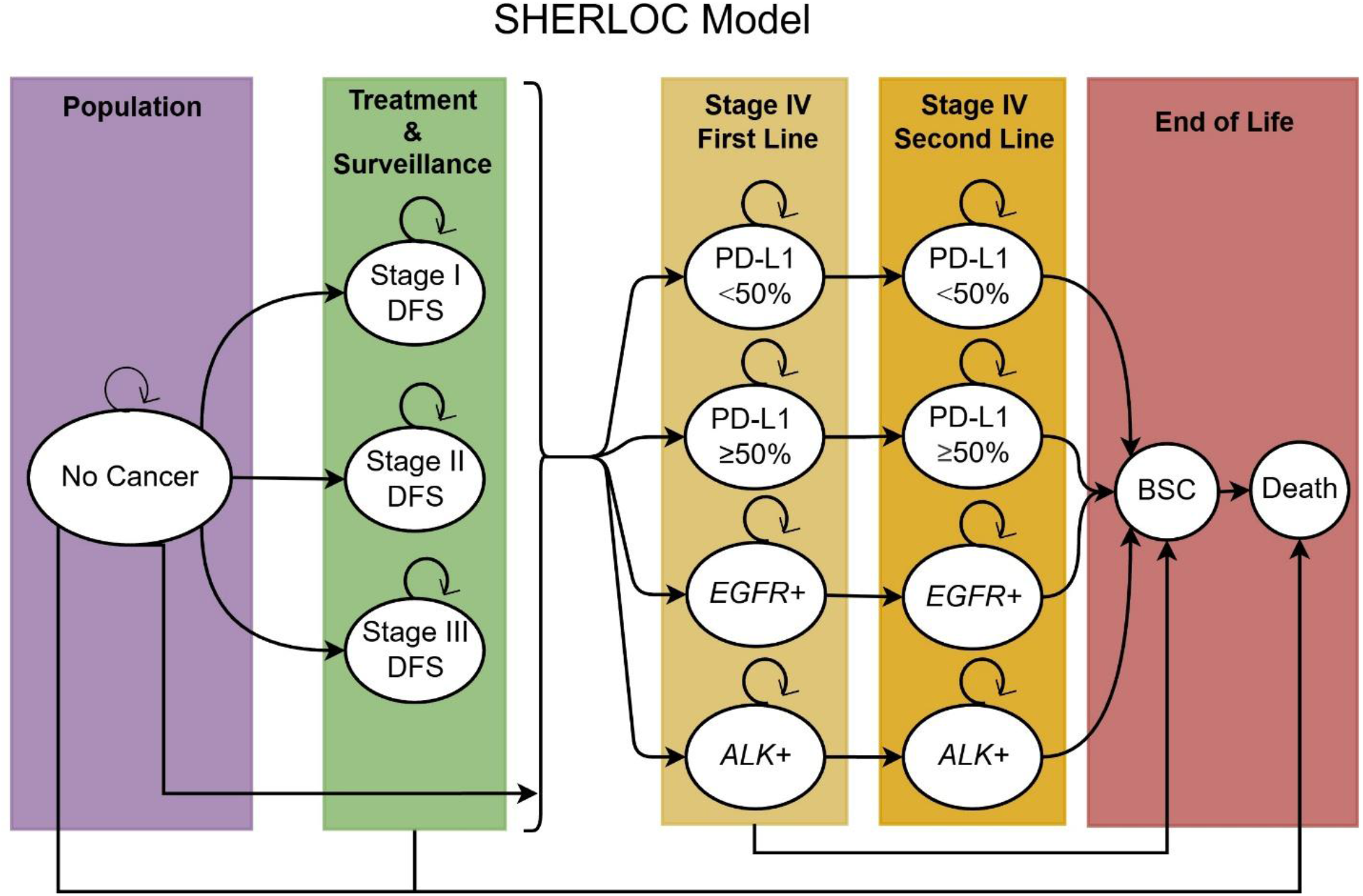
SHERLOC Model Structure DFS=Disease Free Survival, BSC=Best Supportive Care, PD-L1= Programmed Death-Ligand 1 expression

### Model Structure & Assumptions

In order to achieve the required granularity to model NSCLC treatment, a large number of states were required (Figure 1). Accordingly, a microsimulation was chosen, as a cohort model would experience “state explosion” due to the requisite number of substates required to track time in state.^25^ Microsimulations simulate individuals, reducing the mathematical complexity of the model while achieving the same level of accuracy.^25^ ^25^

The disease pathway begins in the no cancer state, where diagnosed individuals are then assigned to a stage (I-IV) and treatment pathway. When assigned to stages I-III, individuals will receive treatment and remain in state until recurrence or death from any other cause. The model assumes all recurrences will be treated as if they are stage IV. Once in stage IV, individuals can receive up to 2 lines of treatment before eventually moving into best supportive care for a cycle and then the absorbing death state. Stage IV is separated into four treatment pathways with PD-L1 <50%, PD-L1 ≥50%, *EGFR*, and *ALK*. While there are a greater number of actionable genomic alterations, only the two most prevalent with publicly funded therapies were chosen as they represent the vast majority of all actionable driver alterations found and treated in NSCLC.^26,27^ In the early stages, treatments are randomly assigned to individuals based on assigned weights (Table 1). Model structure is designed to allow for the assessment of interventions in the first - line stage IV setting for each of the previously noted four treatment pathways. While the model applies a set of transition probabilities for each early stage (I-III) without stratification for the individual treatment categories, the model supports future stratification if assessment of an intervention in the early-stage setting is required. Throughout the model development process, model design, choice of inputs, and modelling of resource usage were performed in line with standards of care guidelines and are described in detail in the supplementary appendix.^26^ The cost of adverse events was applied based on published trial data for adverse events of grade 3 or 4 with a 10% or greater prevalence.^5,21,22,28^

**Table 1:** Model Parameters.

| Parameter | Unit | Cost | SD | Distribution | Source |
|---|---|---|---|---|---|
| **Costs<sup>a</sup>** |  |  |  |  |  |
| Osimertinib: | 28 Day Cycle: 80mg/day | 9,020.00 | ±20% | Gamma | 7 |
| Alectinib: | 28 Day Cycle: 1,200mg/day | 9,918.00 | ±20% | Gamma | 39 |
| Lorlatinib: | 28 Day Cycle: 100mg/day | 8,982.00 | ±20% | Gamma | 38 |
| Pembrolizumab: | 28 Day Cycle: 200mg/3 weeks | 11,733.00 | ±20% | Gamma | 37 |
| Durvalumab: | 28 Day Cycle: 1,500mg/4 weeks | 11,733.00 | ±20% | Gamma | 36 |
| Atezolizumab: | 28 Day Cycle: 1,680mg/4 weeks IV infusion | 9,485.00 | ±20% | Gamma | 35 |
| Pemetrexed: | 28 Day Cycle: 500 mg/m2/3 weeks | 600.00 | ±20% | Gamma | 8 |
| Carboplatin: | 28 Day Cycle: AUC 5 mg/mL/3 weeks | 1,313.20 | ±20% | Gamma | 8 |
| Cisplatin: | 28 Day Cycle: 75 mg/m2/3 weeks | 540.00 | ±20% | Gamma | 8 |
| Docetaxel: | 28 Day Cycle: 100 mg/m2/3 weeks | 2,837.00 | ±20% | Gamma | 41 |
| Nivolumab: | 28 Day Cycle: 3 mg/kg to 240 mg/2 weeks | 9,387.00 | ±20% | Gamma | 40 |
| Amivantamab plus Lazertinib | 28 Day Cycle: Amivantamab: Body weight < 80 kg: 1,050 mg weekly for 4 weeks, then every 2 weeks Body weight ≥ 80 kg: 1,400 mg weekly for 4 weeks, then every 2 weeks Lazertinib: 240 mg once daily | 12,135.00 | ±20% | Gamma | 8 |
| Consultation | Complete assessment | 207.17 | ±20% | Gamma | 42 |
| Office Visit | Routine follow-up | 49.72 | ±20% | Gamma | 42 |
| CT CAP with contrast | Single scan | 1,275.00 | ±20% | Gamma | 45 |
| MRI brain | Single scan | 730.00 | ±20% | Gamma | 44 |
| PET Body | Single scan | 999.00 | ±20% | Gamma | 45 |
| Electrolytes | Per test | 6.53 | ±20% | Gamma | 42 |
| Liver | Per test | 53.57 | ±20% | Gamma | 42 |
| Renal | Per test | 11.65 | ±20% | Gamma | 42 |
| Hematology | Per test | 18.96 | ±20% | Gamma | 42 |
| NGS | Per test | 1,306.75 | ±20% | Gamma | 46 |
| Bronchoscopy | Per intervention | 2856.77 | ±20% | Gamma | 42 & AHS Finance |
| Endoscopic lung resection | Per surgery | 17,079.85 | ±20% | Gamma | 48 |
| Radiation Stage I | 10 Fractions | 8,112.16 | 844110 | Gamma | AHS Finance |
| Radiation Stage II | 10 Fractions | 8,112.16 | 2125660 | Gamma | AHS Finance |
| Radiation Stage III | 30 Fractions | 24,336.49 | 5190353 | Gamma | AHS Finance |
| Hospitalization | Per Stay | 9959.97 | ±20% | Gamma | 43 |
| Best Supportive Care | Per Month | 5772.61 | ±20% | Gamma | 47 |
| **Utilities** |  |  |  |  |  |
| Stage I- Surgery | Quality Adjusted Life Year | 0.82 | 0.16 | Beta | 49 |
| Stage I- Radiation | Quality Adjusted Life Year | 0.76 | 0.2 | Beta | 49 |
| Stage I DFS | Quality Adjusted Life Year | 0.82 | 0.17 | Beta | 49 |
| Stage II - Surgery plus Chemotherapy | Quality Adjusted Life Year | 0.77 | 0.18 | Beta | 49 |
| Stage II - Radiation | Quality Adjusted Life Year | 0.76 | 0.19 | Beta | 49 |
| Stage II DFS | Quality Adjusted Life Year | 0.8 | 0.17 | Beta | 49 |
| Stage III - Chemotherapy plus Radiation | Quality Adjusted Life Year | 0.76 | 0.18 | Beta | 49 |
| Stage III - Surgery plus Chemotherapy | Quality Adjusted Life Year | 0.78 | 0.18 | Beta | 49 |
| Stage III DFS | Quality Adjusted Life Year | 0.79 | 0.19 | Beta | 49 |
| Stage IV: PD-L1 >50% | Quality Adjusted Life Year | 0.71 | 0.24 | Beta | 52 |
| Stage IV PD-L1: <50% | Quality Adjusted Life Year | 0.71 | 0.24 | Beta | 52 |
| Stage IV Actionable | Quality Adjusted Life Year | 0.8 | 0.16 | Beta | 52 |
| IV PD-L1: >50 2L | Quality Adjusted Life Year | 0.67 | 0.2 | Beta | 52 |
| IV PD-L1 <50% 2L | Quality Adjusted Life Year | 0.67 | 0.2 | Beta | 52 |
| IV Actionable 2L | Quality Adjusted Life Year | 0.64 | 0.13 | Beta | 53 |
| BSC | Quality Adjusted Life Year | 0.473 | 0.02 | Beta | 50 |
| FLAURA2 Disutility | Quality Adjusted Life Year | 0.039 | ±20% | Beta | 11 |
| MARIPOSA Disutility | Quality Adjusted Life Year | 0.019 | ±20% | Beta | 11 |
| **Treatment Weights** |  |  |  |  |  |
| Stage I - Surgery | Probability | 0.52349 | - | Fixed | Glans-Look Sample |
| Stage I - Radiation | Probability | 0.47651 | - | Fixed | Glans-Look Sample |
| Stage II - Neo-Adjuvant | Probability | 0.20244 | - | Fixed | Glans-Look Sample & and Expert Opinion |
| Stage II - Radiation | Probability | 0.3252 | - | Fixed | Glans-Look Sample & and Expert Opinion |
| Stage II - Adjuvant | Probability | 0.47236 | - | Fixed | Glans-Look Sample & and Expert Opinion |
| Stage III - Neo-Adjuvant | Probability | 0.09 | - | Fixed | Expert Opinion |
| Stage III - Unresectable | Probability | 0.7 | - | Fixed | Expert Opinion |
| Stage III - Adjuvant | Probability | 0.21 | - | Fixed | Expert Opinion |
| **Cohort Characteristics: Glans-Look Sample n=868** |  |  |  |  |  |
| Stage I - Diagnosis | Probability | 0.16 | - | Fixed | Glans-Look Sample |
| Stage II - Diagnosis | Probability | 0.16 | - | Fixed | Glans-Look Sample |
| Stage III - Diagnosis | Probability | 0.06 | - | Fixed | Glans-Look Sample |
| Stage IV - Diagnosis | Probability | 0.62 | - | Fixed | Glans-Look Sample |
| EGFR | Probability | 0.2501 | - | Fixed | Glans-Look Sample |
| ALK | Probability | 0.1374 | - | Fixed | Glans-Look Sample |
| Stage IV: PD-L1 >50% | Probability | 0.2959 | - | Fixed | Glans-Look Sample |
| Stage IV PD-L1: <50% | Probability | 0.3165 | - | Fixed | Glans-Look Sample |
| **Cohort Characteristics: Alberta** |  |  |  |  |  |
| Stage I - Diagnosis | Probability | 0.26 | - | Fixed | 32 |
| Stage II - Diagnosis | Probability | 0.07 | - | Fixed | 32 |
| Stage III - Diagnosis | Probability | 0.17 | - | Fixed | 32 |
| Stage IV - Diagnosis | Probability | 0.49 | - | Fixed | 32 |
| ALK | Probability | 0.0378 | - | Fixed | 33 |
| EGFR | Probability | 0.2079 | - | Fixed | 33 |
| Stage IV: PD-L1 >50% | Probability | 0.32435 | - | Fixed | 27 |
| Stage IV PD-L1: <50% | Probability | 0.42995 | - | Fixed | 27 |
| **Hazard Ratios** |  |  |  |  |  |
| FLAURA2 TTNT | Hazard Ratio | 0.73 | 0.07398 | Joint Multivariate LogNormal | 21 |
| FLARUA2 OS | Hazard Ratio | 0.77 | 0.08929 | Joint Multivariate LogNormal | 21 |
| MARIPOSA TTNT | Hazard Ratio | 0.76 | 0.06633 | Joint Multivariate LogNormal | 22 |
| MARIPOSA OS | Hazard Ratio | 0.75 | 0.07908 | Joint Multivariate LogNormal | 22 |
| EGFR Endpoint Correlation | Correlation | 0.641 | - | Fixed | 76 |

DFS=Disease Free Survival, PD-L1≥/<50%= Programmed Death-Ligand 1 expression
<sup>a</sup>: includes only drug acquisition costs.

### Programming & Performance

All modelling and statistical analyses were done in R (version 4.3.2). Initial coding was assisted by 2 published modelling tutorials and another open source model, the ALL Policy model in R.^25,29,30^ 10,000 first-order simulations were chosen as the Monte Carlo Squared Error was ∼1% of the mean estimate of cost per case and 5% in the rarest subgroup, and the model appeared to have visually converged (Supplementary Figure 1). Parallel processing is utilized in both the first-order (deterministic) and second-order (probabilistic) models. With a 12th Gen Intel(R) Core(TM) i7-12700H with 16GB of RAM, utilizing 15 cores, 10,000 first-order simulations took 18.18 seconds, and 500 second-order simulations took 5,032seconds. Model code and inputs can be found at [https://github.com/DCGriffin215/SHERLOC-Model]

### Transition Probabilities

Transition probabilities are estimated from a de-identified real-world cohort of lung cancer patients in Alberta from the Glans-Look Lung Cancer Research (GLR) database with diagnoses from 2014-2019 for stage I-III cases and diagnoses from 2018-2023 for all stage IV cases. A diagram detailing chart review inclusion and exclusion is in Supplementary Figure 2, with baseline characteristics including age at diagnosis and sex for all cases, with ECOG performance categorized into 0-1 & ≥2 and smoking status recorded in the stage IV group, shown in Supplementary Table 1. Time-to-events were collected with multistate modelling approaches in mind, where events for early-stage patients were characterized as death prior to recurrence or at recurrence.^31^ Events for late-stage patients were defined as death or time to next treatment, with 2 lines of treatment modelled due to the limited sample size of patients reaching 3rd line treatment. Parametric survival models were fit for state transitions, with Weibull, Log-normal, Log-logistic, Exponential, Gompertz, Gamma, and Generalized Gamma distributions assessed. Fit for extrapolation was determined through the lowest Akaike’s Information Criterion (AIC) and Bayesian Information Criterion (BIC), along with visual inspection for plausibility. Covariates assessed were age and sex for stage I-III transitions, and age, sex, and performance status assessed for stage IV transitions, with covariates selected based on model/estimate stability and a rule of ∼10-15 events per degree of freedom. A full list of selected distributions and covariates can be found in Supplementary Table 2, along with the parametric distribution parameters, coefficients, and confidence intervals along with alternate distributions. All parametric extrapolations, AICs, and BIC for all 18 modelled survival curves can be found in Supplementary Figures 3-20.

### Simulated Population

The SHERLOC model simulates a heterogeneous population including age, sex and performance status. Initial stage distribution of cases is from the 2024 Report on Cancer Statistics in Alberta, and the treatment pathway is determined by frequency in the real-world cohort (Table 1).^32^ Distribution of histological subtypes in stage IV was sourced from literature so as to best represent the Canadian population and is assigned to individuals at the start of each individuals simulation.^27,33^ Individuals are then assigned ages and sex according to initial stage/mutation by truncated normal distributions and random sampling, respectively. For stage IV cases, ECOG status is assigned by logistic regressions and calibrated probability thresholds predicting ECOG according to age. ECOG status is predicted in early-stage cases only when they recur, and their age has been advanced according to their time in state. The resulting simulated population is shown in Supplementary Table 1.

### Model Inputs

Treatment costing and utilities in all states are time-dependent to best capture the costs and health utility of NSCLC. The timing of the monthly costing of therapies was done in line with respective treatment guidelines, with resource use modelled to best resemble standards of care currently in the Alberta Healthcare System.^26,34–48^ Health utilities are collected from the literature, with individuals receiving appropriate health utilities for their respective treatment to account for the utility decrement of adverse events (Table 1).^49–53^ The model utilizes a large number of parameters so that costs can be easily altered for use in other healthcare systems.

### Model Validation

In line with the ISPOR-SMDM Modeling Good Research Practices Report, Internal, face, and external validation were undertaken to assess model validity and comparability to real-life practice.^54^ Internal validation was performed by generating Kaplan-Meier curves and comparing the models’ simulated population’s overall survival at time points (1 year, 3 years, 5 years) to the GLR cohort sample used to inform transition probabilities, along with subgroups such as initial stage and mutation-specific survival estimates, to assess whether structural assumptions have introduced unacceptable error. To assess error, the mean absolute difference (MAE) and mean absolute percentage error (MAPE) were calculated with stage distribution and mutation prevalence for stage IV set to match the Glans-Look Cohort sample characteristics (Table 1). A coefficient of determination (R^2^) statistic was estimated through a linear regression of observed and predicted probabilities to assess the model’s goodness of fit. Both internal and external survival prediction were presented as scatter plots, with external validation additionally presented as line plots of predicted and averaged observed survival at time points. Face validity was assessed through black box testing, multi-author consensus on model code, and utilizing clinician advice throughout the development process.

External validation was performed by comparing model results using stage distributions from the 2024 Canadian cancer statistics report and the reported mutational profile for glans -look (Table 1).^32,33^ Reported survival statistics for 1, 2, 3, and 5 years from publicly available data from comparable health systems in British Columbia (BC), Ontario, Alberta, Canada-wide, and the American Surveillance Epidemiology and End Results (SEER) programs’ most recent reported results for NSCLC as a total disease.^55–59^ In the cases of Alberta, BC, Ontario, and Canada-wide, overall survival is reported for lung cancer total, including SCLC, which currently has poorer survival outcomes; as such, the percent difference between NSCLC and lung cancer total in the SEER cohort was used to adjust reported estimates (Supplementary Table 2).^58^ The estimated difference between lung cancer and NSCLC was calculated by estimating NSCLC survival at 1,3 and 5 years by calculating the weighted average for each of the NSCLC subtypes and then calculating the percent improvement at each yearly time point. Stage-specific survival estimates were also compared against the SEER program and available literature.^58,60,61^ As SEER reports local, regional, and distant survival, stage IIA vs IIB was a 22.6%-77.4% combination of local and regional estimates due to stage IIA being defined as local and stage IIB being regional.^62^ Genomic alteration-specific survival at stage IV was compared against published survival results in real-world observational studies internationally and trial results when no real-world studies were available.^6,63–75^ A total of 75 observations were collected for external validation, with all internal and external data points reported in Supplementary Table 2. Additionally, the reported average cost per case, LYs, and QALYs will be reported for the overall model and subgroups by initial stage and histology for stage IV. A case study was further performed, and the results were compared to CDA-AMC evaluations as this model version was designed for the Canadian setting for further validation.^8^

### Case study: Cost-Effectiveness Evaluation of FLAURA2 and MARIPOSA Trials

The recently published FLAURA2 and MARIPOSA trial results were evaluated to demonstrate the usefulness of the model for assessing interventions and novel technologies in NSCLC treatment and prevention. In the FLAURA2 trial, the current standard of care, osimertinib monotherapy was compared to osimertinib plus platinum-pemetrexed chemotherapy as first-line treatment for *EGFR*-alteration positive advanced NSCLC.^17^ In the MARIPOSA trial, osimertinib monotherapy was compared to amivantamab plus lazertinib for first-line treatment of *EGFR*-alteration positive (*EGFR*+) advanced NSCLC.^18^ Overall survival and time to next treatment hazard ratios were selected from both trials to be applied to the *EGFR*+ first-line overall survival and time to next treatment survival curves. Publication and supplementary materials, while limited in information on post-progression effects, did not indicate any significant effect on post-progression survival beyond the effect from first-line treatment, given the difference between median survivals and hazard ratios remaining unchanged between TTNT1/TTNT2 in the FLAURA2 trial and relatively similar hazard ratios for PFS and PFS2 in the MARIPOSA trial, so the second-line survival curve was not altered.^21,22,76^ Costing was done in accordance with the trial methods, with the first 3 cycles receiving osimertinib plus platinum (cisplatin)-pemetrexed followed by maintenance osimertinib plus pemetrexed until progression or death for the FLAURA2 scenario, with second-line receiving 50% platinum chemotherapy and 50% receiving docetaxel.^21^ Amivantamab plus lazertinib was used until progression or death in the MARIPOSA scenario.^22^ Hazard ratios were applied to monthly transition rates and assumed a constant treatment effect, given both trials found no violation of the proportional hazards assumption and CDA-AMC utilized PH-based parametric models for extrapolations.^21,22,76,77^

The trials indicated an increase in adverse events for both trials, with the NICE assessment utilizing treatment-specific utilities in a scenario.^11^ As such, individuals who receive the FLAURA2 treatment receive a disutility of 0.039 compared to osimertinib monotherapy, as indicated in NICE documents, and a disutility of 0.019 compared to osimertinib monotherapy for MARIPOSA, as trial data indicated dermatological adverse events could be mitigated.^11,78^ Results of the evaluation were incremental costs, incremental QALYs, Incremental Cost - Effectiveness Ratio (ICER), and Incremental Net Monetary Benefit (INMB). Results were evaluated with willingness to pay ratios of $50,000 and $100,000, as is standard for Canadian Health Technology Appraisals.^12^ Reported results are means for incremental costs and QALYs for patients with *EGFR*+ disease, ICER from those means, as ICERs are susceptible to being skewed. Deterministic results are reported for the *EGFR*+ subcohort and compared to evaluations performed by CDA-AMC and a previous evaluation based on interim evidence. Subgroup analyses in both trials showed no significant difference (intersecting hazard-ratio confidence intervals) between subgroups based on sex, race, *EGFR*-alteration type, CNS involvement, or performance score.^21,22^ Therefore, scenario analyses based on subgroups were not conducted.

### Uncertainty

To account for uncertainty, 500 pre-sampled second-order Monte Carlo simulations were conducted with every parameter assigned a parametric distribution informed by either its 95% confidence interval or a standard ±20% from the estimate if a confidence interval or standard deviation is not available and uncertainty is present. A scenario utilizing a ±50% estimate was also utilized. Parametric distribution parameters and coefficients were pre-sampled utilizing the regression covariance matrices to sample parameters from joint multivariate distributions. A scenario was conducted with an alternate model utilizing parametric distributions where AIC and BIC disagreed or an alternate distribution was selected. The selected hazard ratios from the FLAURA2 and MARIPOSA trials were also pre-sampled 500 times from a joint multivariate distribution and applied to the first-line OS and TTNT curves to account for uncertainty in trial comparability to real-world treatment, with an endpoint correlation of 0.641 from a meta-analysis of PFS and OS endpoints for EGFR-TKIs.^79^ The disutilities for osimertinib plus chemotherapy and amivantimab plus lazertinib treatment were sampled with a ±20/50% estimate to maintain the underlying relationship between treatments, but still account for uncertainty. Results of the second-order simulations will be reported on a cost-effectiveness acceptability plane and summarized with cost-effectiveness acceptability curves.

## Results

### Mean Estimates for NSCLC

Estimated mean outcomes and 95% Confidence intervals in costs, QALYs, and LYs for the SHERLOC models are presented in Table 2, with the mean cost per case estimated to be $205,470 (95% CI: $200,950-$209,990) with estimated average QALYs per case of 3.39 (95% CI: 3.31-3.46) and 4.43 (95% CI: 4.34-4.53) LYs. The alternate model reduced QALYs in stage II and III, increased costs and QALYs in the *EGFR+* subcohort, and reduced costs and QALYs in the ALK-alteration positive subcohort (Supplementary Table 3).

**Table 2:**
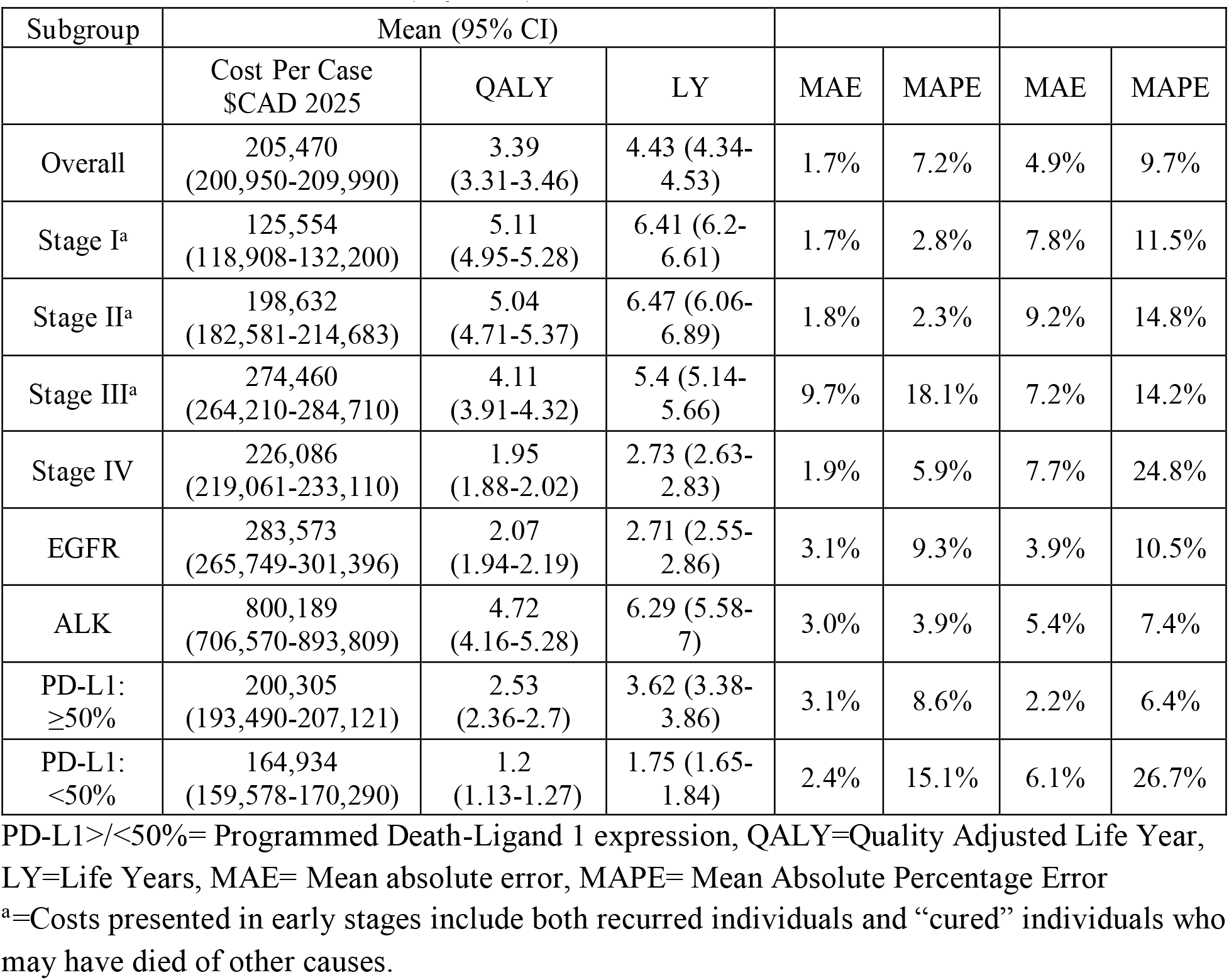
Discounted Mean Cost, QALYs, and LYs Per Case and Validation Error.

### Model Validation

Comparison of internal estimated probabilities with the GLR Cohort for overall survival, stage-specific, and mutation-specific survival was found to have an overall average MAE of 3.1% and an MAPE of 8.1% with an R^2^ of 0.9721, shown in Figure 2 and Table 2. Comparison to external healthcare systems found that the models estimated survival for total NSCLC had an MAE of 4.9% and an MAPE of 9.7%. Stage-specific survival predictions had an MAE of 8% and an MAPE of 16.3%, (Supplementary Figure 21 & Table 2). Genomic alteration-specific survival in stage IV had an average MAE of 4.4% and an MAPE of 12.7%, with PD-L1 ≥50% being the best fitting (MAE: 2.2%, MAPE: 6.4%) and PD-L1 <50% being the worst (MAE: 6.1%, MAPE: 26.7%) (Supplementary Figure 21 & Table 2). The overall average external MAE was 6%, and an average MAPE of 14%, with a calculated adjusted R^2^ of 0.8795 (Figure 2 & Table 2). The alternate model slightly improved average internal MAPE (7.5%) and worsened MAE (3.2%) (Supplementary Table 3 & Supplementary Figure 22). Externally, the alternate distributions reduced error in the *EGFR+* and ALK-alteration positive groups and increased error in stage II and stage III simulated survival (Supplementary Table 3 & Supplementary Figure 22).

**Figure 2:**
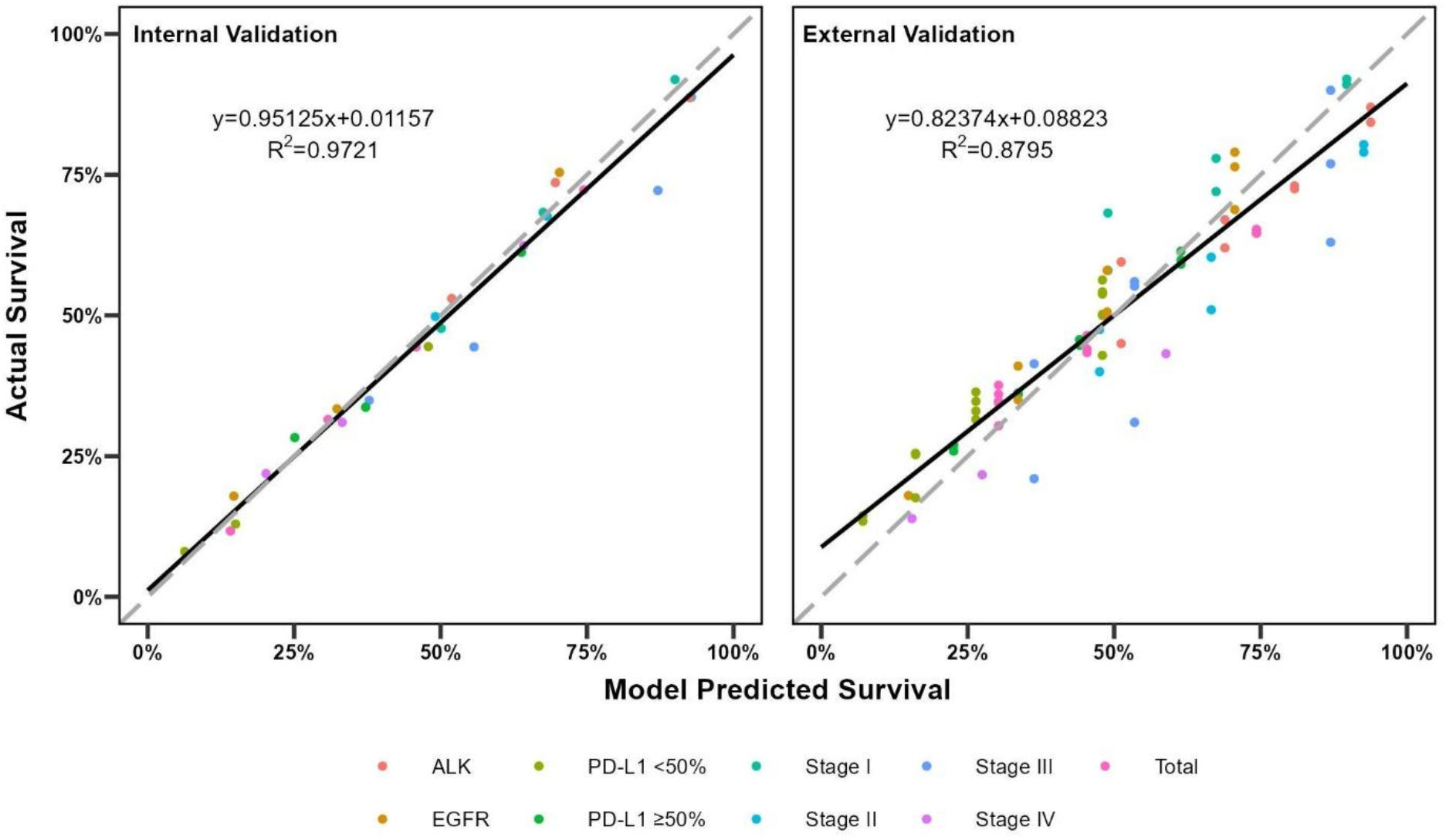
Scatter plots and fitted linear regression results for internal validation and external validation of survival probabilities against the GLR cohort and observational results, where the black line represents the fitted model and the grey dashed line represents a 45-degree reference line.

### Case Study Results

The Incremental costs for the *EGFR*+ subcohort in the base case were $110,525 (95% CI: $106,969-$114,082) for FLAURA2 and $186,855 (95% CI: $181,060-$192,651) for MARIPOSA, Incremental QALYs were 0.413 (95% CI: 0.389-0.436) for FLAURA2 and 0.483 (95% CI: 0.461-0.504) for MARIPOSA (Table 3). The ICERs comparing each to osimertinib monotherapy were $267,908 and $387,223 per QALY gained, osimertinib with platinum-pemetrexed chemotherapy extendedly dominated amivantamab plus lazertinib in 85.8% of simulations (Table 3 and Figure 3). The increased uncertainty assumption resulted in FLAURA2 extendedly dominating MARIPOSA in 70.4% of simulations (Table 3 and Supplementary Figure 24. The alternate model with a loglogistic 1L-BSC curve increased incremental costs $138,975 (95% CI: $134,559-$143,391) & $221,477 (95% CI: $214,911-$228,043) and QALYs 0.576 (95% CI: 0.546-0.605) & 0.627 (95% CI: 0.601-0.653). ICERs only slightly decreased to $241,394 and $353,086 per QALY gained (Table 3 & Supplementary Figure 24). The findings of the cost-effectiveness analysis are relatively in line with the findings of Canada’s Drug Agency with neither being likely to be cost-effective at a $50,000 or $100,000 willingness-to-pay ratio (Table 3).^8^ In CDA-AMC’s reanalysis, partition survival models were utilized, assuming equal efficacy, resulting in probabilistic incremental QALYs that were identical with probabilistically 0.66 QALYs gained for each and deterministic 0.49 QALYs gained, indicating a large skew in the probabilistic analysis. Incremental costs were $119,721 for FLAURA2 and $201,023 for MARIPOSA against the standard of care.^8^

**Figure 3:**
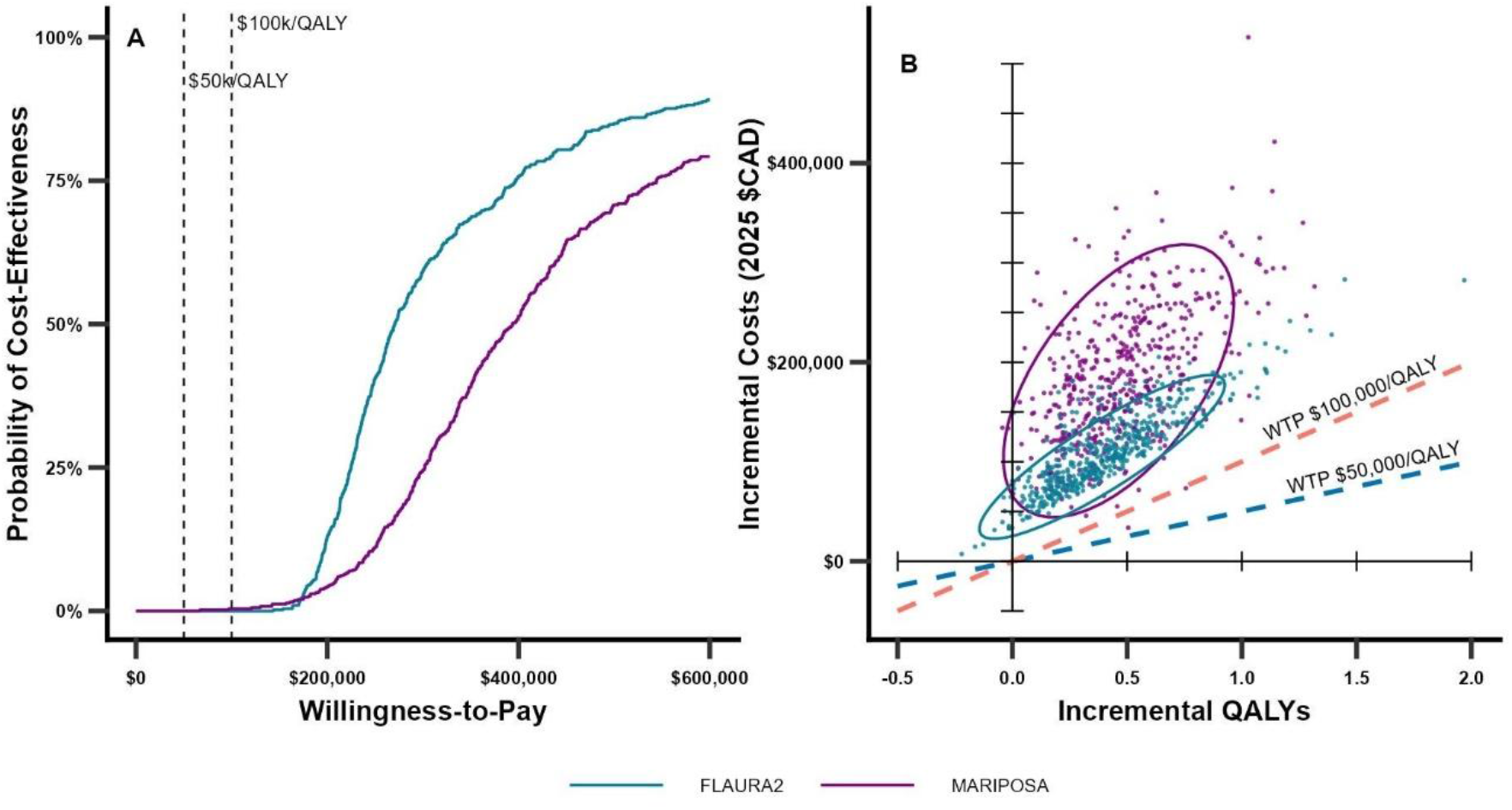
Cost-effectiveness acceptability curves (**A**) and cost-effectiveness acceptability plane and 95% confidence ellipse (**B**) with osimertinib plus platinum pemetrexed for FLAURA2 (Blue) and amivantamab plus lazertinib for MARIPOSA (Purple) against osimertinib monotherapy.

**Table 3:** Cost-Effectiveness Results For FLAURA2 and MARIPOSA against Osimertinib Monotherapy.

| Cohort | Trial | Incremental Mean Cost Per Case (95% CI) \$CAD | Incremental mean QALYs (95% CI) | Incremental Cost-Effectiveness Ratio \$CAD | Probability of Cost-Effectiveness at Willingness to Pay Ratios: |  | Incremental Net Monetary Benefit (INMB) \$50,000 WTP |
|---|---|---|---|---|---|---|---|
|  |  |  |  |  | \$50,000 | \$100,000 |  |
| SHERLOC Model: EGFR Subcohort Base Case | FLAURA2 vs. SOC | **110,525** (106,969-114,082) | **0.413** (0.389-0.436) | **267,908** per QALY | 0% | 0% | **-89,897** (-92513 : -87282) |
|  | MARIPOSA vs. SOC | **186,855** (181,060-192,651) | **0.483** (0.461-0.504) | **387,223** per QALY | 0% | 0.40% | **-162,727** **(**-168,002 : -157,453**)** |
|  | MARIPOSA vs. FLAURA2 | **76,330** (69,728-82,932) | **0.07** (0.044-0.096) | **1,090,396** per QALY | **-** | **-** | **-72,830** (-78,601 : -67,058) |
| SHERLOC Model: EGFR Subcohort 50% uncertainty | FLAURA2 vs. SOC | **110,590** (106,648-114,530) | **0.412** (0.389-0.436) | **268,248** per QALY | 0% | 0% | **-89,976** (-93,075 : -86,878) |
|  | MARIPOSA vs. SOC | **185,048** (173,491-196,606) | **0.482** (0.461-0.503) | **383,700** per QALY | 8% | 12% | **-160,935** **(**-172,240 : -149,630**)** |
|  | MARIPOSA vs. FLAURA2 | **74,459** (61,728-87,190) | **0.07** (0.044-0.096) | **1,063,543** per QALY | **-** | **-** | **-70,959** (-83,248 -58,670) |
| SHERLOC Model: EGFR Subcohort Alternate Model | FLAURA2 vs. SOC | **138,975** (134,559-143,391) | **0.576** (0.546-0.605) | **241,394** per QALY | 0% | 0% | **-110,189** (-113,400 : -106,978) |
|  | MARIPOSA vs. SOC | **221,477** (214,911-228,043) | **0.627** (0.601-0.653) | **353,086** per QALY | 0% | 0% | **-190,114** **(**-196,043 : -184,184**)** |
|  | MARIPOSA vs. FLAURA2 | **82,502** (74,939-90,064) | **0.052** (0.019-0.084) | **1,600,682** per QALY | **-** | **-** | **-79,924** (-86,447 -73,402) |
| CDA | FLAURA2: Deterministic | 123,145 | 0.49 | 250,266 | NR | NR | NR |
|  | MARIPOSA: Deterministic | 203,322 | 0.49 | 414,943 | NR | NR | NR |
|  | FLAURA2: Probabilistic | 119,721 | 0.66 | 181,395 | NR | NR | NR |
|  | MARIPOSA: Probabilistic | 201,023 | 0.66 | 304,580 | NR | NR | NR |

QALY=Quality-Adjusted Life Years, Costs presented in 2025 \$CAD, NR=Not Reported

## Discussion

In this publication, we aimed to develop and validate an open-source disease model for NSCLC that is capable of assessing both preventative interventions and new oncology therapies. The model was validated internally to assess if assumptions had introduced error, and externally to assess if the model is valid against NSCLC in the Canadian health system.

Results from the model’s validation, with an average internal MAE of 3.1%, an MAPE of 8.1%, and an R^2^ of 0.9721, along with an external average MAE of 6% and MAPE of 14% with an R^2^ of 0.8795, indicate that model assumptions have acceptable levels of error internally and are in line with complex models validated in similar manners. To put these statistics into context, the iTEN model, a discrete event simulation model for advanced non-small cell lung cancer, was externally validated with an average MAPE of 30.6% (Range: 49.3%-11.6%) with an R^2^ of 0.96.^80^ txSim, a model for advanced lung cancer, had an average MAE of 5% (Range: 1.6%-11.7%) and MAPE of 21.6% (Range: 9.1%-32.6%).^81^ Model validation in this respect is more common in diabetes, with models such as the Cardiff model reporting an average external MAPE of 37.7% (Range: 20.4%-89.9%) and an R^2^ of 0.87.^82^ More recent examples in diabetes reported an internal mean absolute deviation of 7.9% and R^2^ of 0.9907 in one case and 0.975 in another.^83,84^ A further study reported external independent MAPEs ranging from 37.2% to 86.1% with respective R^2^ of 0.905 and 0.81.^85^ The worst MAPE value (PD-L1 <50%: 26.7%) in the model is lower than many percent error values of these comparators. Further, due to PD-L1 <50% having the worst survival, 5-year time points could only be found in trials whose reported survival is likely optimistic. Stage IV (MAPE: 24.8%) is only compared to SEER data, as survival at stage IV is rarely reported for NSCLC, and it is more commonly broken down by subgroup.

When compared to the CDA-AMC re-analysis, the results are similar, with this analysis finding that both trials result in similar survival gains, like the CDA-AMC re-analysis, while the incremental results are slightly smaller than the CDA results.^8^ The skewed deterministic vs probabilistic results in the CDA-AMC evaluation, where incremental QALY gains increase by 35%, inhibits a proper comparison without more information as the cause is unknown. While both analyses ultimately find the trials to be not cost-effective and are largely in agreement, this further highlights the differences that can occur in modelling the same decision problem with different assumptions, structures, and evidence, while highlighting the need for transparency in modelling.^8^ While the CDA-AMC analysis finds larger incremental QALYs and LYs, the incremental increases are proportionally similar (CDA-AMC: 24%, MARIPOSA Modelled: 21.5%), considering the higher base QALYs for osimertinib therapy of 2.75 vs 2.08 (Table 2).^8^ This is in line with findings that trial populations tend not to reflect real-world heterogeneity, indicating this analysis likely better reflects real-world implementation (Supplementary Figure 3 & Table 2).^86^

The ISPOR-SMDM Modeling Good Research Practices Report recommends internal, external, face, and cross-validation; however, cross-validation was not performed in this analysis.^54^ This is due to OncoSim-Lung being the only model in the Canadian context that could potentially be comparable to this model. Because of the outdated nature of some of its parameters concerning survival and costs, along with the model’s design for preventative interventions, cross-validation at this time was not performed.^14,15^ When OncoSim-Lung is updated, future research could cross-validate the SHERLOC model through the assessment of screening programs in Canada.

The model is notably limited in some respects. While external validation of survival results in comparable health systems in Canada and internationally assists with generalizability, healthcare systems are independently run with differing standards of care and differing speeds of adoption of new technologies, along with independently negotiating drug prices with little to no transparency on negotiated pricing. Research supports the comparability of survival estimates in advanced NSCLC between Canada and the US, but the previously noted limitations around costs and speed of adoption of new technologies apply.^87^ This is mitigated in the model through second-order simulations including uncertainty in prices, utilities, and transition probabilities, but is nonetheless worth noting. Additionally, the adjustment of reported overall lung cancer survival estimates to NSCLC using SEER data potentially underestimates the differences between NSCLC and LC in the Canadian population due to historically delayed adoption of new standards of care.^88^ The modelling of only two lines of treatment in stage IV disease may result in bias in the estimated costs that could potentially bias the estimated cost of treatment; however, a brief quantitative analysis found minimal potential cost bias given the available third-line data as is detailed in the Supplementary Appendix. A mitigation for many of these limitations is the model being open source, with model inputs easily changed to reflect costs or health utilities in an external setting. The use of a PFS-OS correlation for sampling uncertainty in the hazard ratios is potentially an imperfect correlation. However, a retrospective TTNT OS correlation in real-world oral TKIs for NSCLC was 0.62, indicating minimal potential bias.^89^

## Conclusion

This study describes the development and validation of the SHERLOC model, with the goal of providing a transparent and validated platform that could be used in the economic evaluation of interventions in NSCLC and lung cancer at large. Validation of the model found that the model performed well when compared to the cohort used to inform transition probabilities and had external performance in line with other published models in lung cancer and diabetes, but cross-validation is still needed to further validate the model. The model then assessed two competing trials in the first-line *EGFR*+ advanced NSCLC setting, finding that neither was likely to be cost-effective at a $50,000 or $100,000 willingness-to-pay ratio and osimertinib plus platinum pemetrexed extendedly dominated amivantamab plus lazertinib.

## Data Availability

Datasets related to chart review data for the current study are not available due to institutional and ethical restrictions.

## Acknowledgments

The authors would like to acknowledge Dr. Doreen Ezeife (University of Calgary, Alberta, Canada) for her contributions in clinical expertise in developing the base model structure and standard of care treatment costing.

## Disclosures

Vishal Navani

Disclosures:

•Consulting – Novotech Pty, Pfizer, Sanofi, Astra Zeneca, EMD Serono, Oncology Education, Sanofi, Janssen, Roche, MSD, Bristol Meyers Squibb, Takeda, Boeringer Ingleheim

•Speaking – Ipsen, Astra Zeneca, MSD, Bristol Meyers Squibb

•Research – Astra Zeneca (Inst), Janssen (Inst)

•Travel – EMD Serono, Pfizer, Sanofi

