## Supplementary Appendix for "Development and Validation of the SHERLOC Model: A Disease Model for Non-Small Cell Lung Cancer"

**Costs:**

Stage I is divided into 2 treatment categories: surgery or radiation. Radiation is divided into the first 2 cycles, and Endoscopic surgery is applied in the first cycle. Once treatment has been completed a cost for surveillance is applied for 5 years following treatment.

Stage II is divided into radiation, neoadjuvant plus surgery, and adjuvant plus surgery. Radiation is again divided into the first 2 months, neoadjuvant receives 2 cycles of Nivolumab then surgery, and adjuvant plus surgery receives surgery and 3 cycles of cisplatin plus pemetrexed from months 1 to 3. 50% of adjuvant plus surgery cases receive 1 year of atezolizumab post chemotherapy. Once treatment has been completed a cost for surveillance is applied for 5 years following treatment.

Stage III is divided into unresectable, neoadjuvant plus surgery, and adjuvant plus surgery. Unresectable receives 2 cycles of radiation and 3 cycles of Cisplatin plus Pemetrexed, then a year of Durvalumab. neoadjuvant receives 2 cycles of Nivolumab then surgery, and adjuvant plus surgery receives surgery and 3 cycles of cisplatin plus pemetrexed from months 1 to 3. 50% of adjuvant plus surgery cases receive 1 year of atezolizumab post-chemotherapy. Once treatment has been completed a cost for surveillance is applied for 5 years following treatment.

EGFR 1L receives Osimertinib till progression or no longer able to tolerate treatment. ALK 1L receives Alectinib till progression or no longer able to tolerate treatment. PD-L1 ≥50% receives Pembrolizumab for 2 years. PD-L1 <50% receives Carboplatin plus Pemetrexed plus Pembrolizumab for 3 cycles and then Pemetrexed plus Pembrolizumab till 2 years total. Once treatment has ceased for PD-L1 cases, costs for surveillance are applied till progression or Death

EGFR 2L receives Carboplatin plus Pemetrexed, ALK receives Lorlatinib, PD-L1 ≥50% receives Carboplatin plus Pemetrexed, PD-L1 <50% receives docetaxel.

Early-stage patients after treatment receive surveillance of 2 CT scans plus an office visit yearly for up to 5 years if the simulated individual remains disease-free, and in the case of late-stage pembrolizumab treatment, once treatment is stopped at 2 years, individuals receive the cost of 4 CT scans plus an office visit per year till progression or death.

Other costs are determined by stage with costs for early stages being costs applied during the duration of treatment, and costs for stage IV being yearly costs. Stage I receives 1x consultation, 2x office visit, 1x PET scan, 1x brain MRI, 2x chemistry, 2x hematology, 1x bronchoscopy. stage II receiving 1x consultation, 5x office visit, 1x CT with contrast, 1x brain MRI, 1x PET scan, 6x chemistry, 6x hematology, 1x bronchoscopy. stage III and IV receiving 1x consultation, 17x office visit, 4x CT with contrast, 1x brain MRI, 1x PET scan, 20x chemistry, 20x hematology, 1x Next Generation Sequencing, 1x bronchoscopy. Stage IV second line receives 17x office visit, 4x CT with contrast, 17x hematology, and Actionable second line receiving 2 extra brain MRI.

Uncertainty is applied to all costs due to differing costs across Canadian healthcare systems and unknown discounts on CDA-AMC list prices.

**Utilities:**

All Utilities are treatment-specific and are applied to their respective treatment in the model. In the case of early stages, once individuals have ceased treatment and are considered to be in surveillance, they receive long-term follow-up health utilities with a lag time for recovery of 3 months for surgery or chemotherapy and 1 month for radiotherapy to account for lag times in recovery of quality of life.

**Quantitative Assessment of Modelling Two Lines of Treatment**

Potential bias of only modelling two lines of treatment for stage IV cases was assessed using the 59 recorded cases in the GLR dataset that reached third-line treatment, totalling ~11% of all stage IV cases. Recommended third line treatment in Alberta is docetaxel or erlotinib, docetaxel was chosen for this assessment as it is the more expensive of the two with an incremental cost of $1,289 per month over carboplatin plus pemetrexed. Median third line survival was 5 months in the data from start of third line therapy to death. The expected increase in costs if third-line was modelled would be approximately $1,289*5*11.2%, summing to $721.84, or a 0.32% increase in the average cost per stage IV case, indicating minimal cost bias due to not modelling third-line treatment.

**Time Horizon**

To assess the choice of a 25-year time horizon, the survival rate at the end of the 25-year simulation was assessed. After 25 years of simulation, mortality in the model was 97%.

**Survival Analysis:**

Survival Analysis was performed examining cause-specific survival curves, with covariates assessed in all curves. Early-stage curves had age and sex assessed as covariates, and late-stage had age, sex, and ECOG score classified into 0-1 & ≥2. Covariates were selected based on events to degrees of freedom along with estimate and model stability. Parametric distributions were ranked according to AIC and BIC and then plotted utilizing the easysurv package to compare projected distributions. In the case of categorical variables like sex and ECOG. Distributions representing unrealistic long-term extrapolations such as cure fractions in stage IV disease or overly pessimistic extrapolations in early-stage disease, were rejected in favour of the second-ranked distribution. In cases of a disagreement between AIC and BIC and the extrapolation of the alternate distribution was reasonable, an alternate model was created using the alternate distributions to assess the effect of differing long-term extrapolations on results. Survival curves shown are presented with kaplan meier curves to display the shape of the recorded survival curve but in the cases of variables like ECOG and sex the curves presented are for each dichotomous value/combination of values and will not line up with the Kaplan meier curve shown. Alternate curves were selected for stage II-Death, Stage II-Stage IV, Stage III-Death, ALK-1L-2L, EGFR 1L-2L.

**Supplementary Table 1:**

|  | **Overall** | **SHERLOC Model Simulated Population** | **Stage I** | **Stage II** | **Stage III** | **EGFR+** | **ALK+** | **PD-L1 <50%** | **PD-L1 >50%** |
| --- | --- | --- | --- | --- | --- | --- | --- | --- | --- |
| **Age: Mean (sd) [range]** | 67.59 (10.78) [25-95] | 67.37 (10.14)  [25-95] | 69.93 (9.82) [34-92] | 71.05 (9.29) [47-95] | 67.22 (9.63) [46-91] | 68.12 (12.33) [30-94] | 58.67 (15.41) [25-87] | 66.39 (9.18) [29-86] | 67.44 (7.54) [51-85] |
| **Sex** | | | | | | | | | |
| **Female** | 55.9% | 56.04% | 56.03% | 54.74% | 56.36% | 58.78% | 52.78% | 53.57% | 58.71% |
| **Male** | 43.7% | 43.96% | 43.97% | 44.53% | 43.64% | 38.93% | 47.22% | 46.43% | 41.29% |
| **missing** | 0.50% | - | - | 0.73% | - | 2.29% | - | - | - |
| **ECOG (Stage IV)** | | | | | | | | | |
| **0-1** | 73.10% | 72.90% | - | - | - | 67.94% | 81.94% | 73.81% | 71.61% |
| **≥2** | 26.90% | 27.10% | - | - | - | 31.30% | 18.06% | 26.19% | 28.39% |
| **missing** | 0.20% | - | - | - | - | 0.76% | - | - | - |
| **Smoking Status (Stage IV)** | | | | | | | | | |
| **Ever Smoker** | 71.20% | - | - | - | - | 44.27% | 26.39% | 89.29% | 94.84% |
| **Never Smoker** | 28% | - | - | - | - | 54.20% | 72.22% | 10.71% | 5.16% |
| **Unkown/missing** | 0.80% | - | - | - | - | 1.53% | 2.78% | - | - |

**Supplementary Table 2:** Chosen Parametric Distributions

| State | Transition | Disribution | Parameter 1: Estimate (95% CI) | Parameter 2: Estimate (95% CI) | Parameter 3: Estimate (95% CI) | Parameter 4: Estimate (95% CI) | Parameter 5: Estimate (95% CI) |
| --- | --- | --- | --- | --- | --- | --- | --- |
| Stage I | Stage I-Death | Exponential | Rate: 2.20e-04 (1.60e-04-3.02e-04) | - | - | - | - |
|  | Stage I-Stage IV | Log Normal | Meanlog: 5.82142 (3.71682-7.92602) | SDlog: 1.50725 (1.23087 -1.84569) | Age: 0.02692 (-0.00352-0.05736) | - | - |
| Stage II | Stage II-Death | Exponential | Rate: 1.41e-04 (9.18e-05-2.16e-04) | - | - | - | - |
|  | Stage II-Stage IV | Generalized Gamma | Mu: 7.73695 (5.54847 9.92543) | Sigma: 1.55968 (1.31083 1.85576) | Q: -1.00333 (-2.01425 0.00758) | Sex: 0.18843 (-0.35752 0.73438) | age: -0.01808 (-0.04720 0.01103) |
| Stage III | Stage III-Death | Log Normal | Meanlog: 8.135 (7.178 9.092) | SDlog: 1.610 (1.032 2.513) | - | - | - |
|  | Stage III-Stage IV | Generalized Gamma | Mu:5.7900 (4.8372 6.7427) | Sigma: 1.5702 (1.1864 2.0783) | Q: -1.1746 (-2.4042 0.0549) | - | - |
| EGFR 1L | 1L-BSC | Exponential | Rate: 5.60e-04 (4.08e-04 7.70e-04) | ECOG: 1.29e+00 7.99e-01 1.78e+00 | - | - | - |
|  | 1L-2L | Exponential | Rate: 4.27e-04 (2.97e-04 6.15e-04) | ECOG: 5.28e-01 (-1.91e-01 1.25e+00) | - | - | - |
| ALK 1L | 1L-BSC | Exponential | Rate: 2.27e-04 (1.32e-04 3.91e-04) | - | - | - | - |
|  | 1L-2L | Log Normal | Meanlog: 7.367 ( 6.837 7.898) | Sdlog: 1.552 (1.164 2.070) | - | - | - |
| PD-L1 >50% | 1L-BSC | Log Normal | Meanlog: 12.4947 (8.7097 16.2797) | SDlog: 2.1812 (1.8361 2.5912) | Sex: -0.7355 (-1.5314 0.0604) | Age: -0.0763 (-0.1302 -0.0224) | ECOG: -1.1022 (-1.9482 -0.2561) |
|  | 1L-2L | Log Normal | Meanlog: 7.5467 (6.9324 8.1609) | SDlog: 1.8762 (1.5035 2.3412) | Sex: 0.1178 (-0.7075 0.9432) | ECOG: -0.0732 (-1.0133 0.8669) | - |
| PD-L1 <50% | 1L-BSC | Log Normal | Meanlog: 5.63747 (3.75866 7.51628) | SDlog: 1.50101 (1.29896 1.73448) | Sex: -0.66758 (-1.18123 -0.15392) | Age: 0.00921 (-0.01874 0.03716) | ECOG: -0.65696 (-1.22058 -0.09335) |
|  | 1L-2L | Log Normal | Meanlog: 4.90821 (3.07140 6.74502) | SDlog: 1.26104 (1.05317 1.50995) | Age: 0.02347 (-0.00412 0.05106) | - | - |
| PD-L1 <50% 2L | 2L-BSC | lognormal | Meanlog: 4.5643 (2.3227 6.8060) | SDlog: 1.2220 (0.9997 1.4937) | Age: 0.0110 (-0.0225 0.0444) | ECOG: 0.5924 (-0.1984 1.3831) | - |
| PD-L1 >50% 2L | 2L-BSC | Log-Logistic | Shape: 1.579 (1.190 2.096) | Scale: 278.263 (200.996 385.232) | - | - | - |
| EGFR 2L | 2L-BSC | Exponential | Shape: 1.53 (1.16 2.02) | Scale: 263.28 (183.72 377.31) | - | - | - |
| ALK 2L | 2L-BSC | Exponential | Rate: 0.000968 (0.000583 0.001605) | - | - | - | - |
| **Alternate** | | | | | | | |
| Stage II | Stage II-Death | Gompertz | Shape: 5.90e-04 (1.15e-04 1.07e-03) | Rate: 7.87e-05 (3.63e-05 1.71e-04) | - | - | - |
|  | Stage II-Stage IV | Log Normal | Meanlog: 8.4343 (6.2345 10.6340) | Sdlog: 1.4749 (1.2379 1.7572) | Sex: 0.2027 (-0.3545 0.7599) | age: -0.0195 (-0.0498 0.0109) | - |
| Stage III | Stage III-Death | Exponental | Rate: 2.31e-04 (1.25e-04 4.30e-04) | - | - | - | - |
| EGFR 1L | 1L-2L | Log Logistic | Shape: 1.256 (1.027 1.536) | Scale: 1333.308 (949.935 1871.404) | ECOG: -1.748 (-2.333 -1.164) | - | - |
| ALK 1L | 1L-2L | Exponental | Rate: 0.000454 (0.000309 0.000666) | - | - | - | - |

**Supplementary Table 3:** Discounted Mean Cost, QALYs, and LYs Per Case and Validation Error in the alternate model

| Subgroup | Mean (95% CI) | | |  | |  | |
| --- | --- | --- | --- | --- | --- | --- | --- |
|  | Cost Per Case $CAD 2025 | QALY | LY | MAE | MAPE | MAE | MAPE |
| Overall | 205,765 (201,297-210,233) | 3.3 (3.22-3.37) | 4.33 (4.23-4.42) | 1.3% | 3.7% | 4.8% | 9.4% |
| Stage I | 125,941 (119,212-132,669) | 5.12 (4.95-5.28) | 6.41 (6.21-6.62) | 1.9% | 3.1% | 7.9% | 11.6% |
| Stage II | 197,437 (182,512-212,363) | 4.26 (4.04-4.47) | 5.51 (5.24-5.78) | 3.4% | 4.3% | 11.8% | 19.5% |
| Stage III | 273,337 (263,216-283,458) | 3.89 (3.71-4.07) | 5.12 (4.88-5.35) | 10.1% | 19.2% | 7.3% | 14.8% |
| Stage IV | 227,037 (220,099-233,975) | 1.96 (1.88-2.03) | 2.74 (2.64-2.85) | 1.7% | 5.2% | 7.7% | 25.7% |
| EGFR | 300,273 (279,723-320,824) | 2.18 (2.04-2.32) | 2.85 (2.67-3.03) | 3.0% | 5.4% | 3.1% | 7.1% |
| ALK | 735,856 (657,304-814,408) | 4.32 (3.85-4.78) | 5.85 (5.23-6.46) | 2.1% | 2.5% | 5.2% | 7.1% |
| PD-L1: ≥50% | 200,305 (193,490-207,121) | 2.53 (2.36-2.7) | 3.62 (3.38-3.86) | 3.1% | 8.6% | 2.2% | 6.4% |
| PD-L1: <50% | 164,934 (159,578-170,290) | 1.2 (1.13-1.27) | 1.75 (1.65-1.84) | 2.4% | 15.1% | 6.1% | 26.7% |

**Supplementary Table 4:** List of Internal and External Probabilities used for validation

| **Internal** | | | |
| --- | --- | --- | --- |
| Total | 1 year | **74.4%** | SHERLOC Model |
|  |  | **74.3%** | Alternate Model |
|  |  | 72.3% | Glans-Look sample |
|  | 3 year | **45.8%** | SHERLOC Model |
|  |  | **46.7%** | Alternate Model |
|  |  | 44.4% | Glans-Look sample |
|  | 5 year | **30.8%** | SHERLOC Model |
|  |  | **31.5%** | Alternate Model |
|  |  | 31.5% | Glans-Look sample |
|  | 10 years | **14.1%** | SHERLOC Model |
|  |  | **12.5%** | Alternate Model |
|  |  | 11.7% | Glans-Look sample |
| StageI | 1 year | **90.0%** | SHERLOC Model |
|  |  | **89.8%** | Alternate Model |
|  |  | 91.9% | Glans-Look sample |
|  | 3 year | **67.5%** | SHERLOC Model |
|  |  | **67.4%** | Alternate Model |
|  |  | 68.3% | Glans-Look sample |
|  | 5 year | **50.1%** | SHERLOC Model |
|  |  | **50.5%** | Alternate Model |
|  |  | 47.7% | Glans-Look sample |
| Stage II | 1 year | **92.8%** | SHERLOC Model |
|  |  | **94.9%** | Alternate Model |
|  |  | 88.8% | Glans-Look sample |
|  | 3 year | **68.2%** | SHERLOC Model |
|  |  | **71.4%** | Alternate Model |
|  |  | 67.6% | Glans-Look sample |
|  | 5 year | **49.1%** | SHERLOC Model |
|  |  | **50.1%** | Alternate Model |
|  |  | 49.8% | Glans-Look sample |
| Stage III | 1 year | **87.1%** | SHERLOC Model |
|  |  | **87.1%** | Alternate Model |
|  |  | 72.2% | Glans-Look sample |
|  | 3 year | **55.7%** | SHERLOC Model |
|  |  | **56.5%** | Alternate Model |
|  |  | 44.4% | Glans-Look sample |
|  | 5 year | **37.8%** | SHERLOC Model |
|  |  | **38.3%** | Alternate Model |
|  |  | 34.9% | Glans-Look sample |
| Stage IV | 1 year | **64.2%** | SHERLOC Model |
|  |  | **63.5%** | Alternate Model |
|  |  | 62.4% | Glans-Look sample |
|  | 3 year | **33.2%** | SHERLOC Model |
|  |  | **33.8%** | Alternate Model |
|  |  | 31.0% | Glans-Look sample |
|  | 5 year | **20.2%** | SHERLOC Model |
|  |  | **20.9%** | Alternate Model |
|  |  | 21.9% | Glans-Look sample |
| EGFR | 1 year | **70.3%** | SHERLOC Model |
|  |  | **67.8%** | Alternate Model |
|  |  | 75.4% | Glans-Look sample |
|  | 3 year | **32.3%** | SHERLOC Model |
|  |  | **33.8%** | Alternate Model |
|  |  | 33.4% | Glans-Look sample |
|  | 5 year | **14.7%** | SHERLOC Model |
|  |  | **17.0%** | Alternate Model |
|  |  | 17.9% | Glans-Look sample |
| ALK | 1 year | **92.5%** | SHERLOC Model |
|  |  | **92.1%** | Alternate Model |
|  |  | 88.7% | Glans-Look sample |
|  | 3 year | **69.6%** | SHERLOC Model |
|  |  | **71.0%** | Alternate Model |
|  |  | 73.6% | Glans-Look sample |
|  | 5 year | **51.9%** | SHERLOC Model |
|  |  | **52.9%** | Alternate Model |
|  |  | 53.0% | Glans-Look sample |
| PD-L1 ≥50% | 1 year | **63.8%** | SHERLOC Model |
|  |  | **63.8%** | Alternate Model |
|  |  | 61.2% | Glans-Look sample |
|  | 3 year | **37.2%** | SHERLOC Model |
|  |  | **37.2%** | Alternate Model |
|  |  | 33.7% | Glans-Look sample |
|  | 5 year | **25.1%** | SHERLOC Model |
|  |  | **25.1%** | Alternate Model |
|  |  | 28.3% | Glans-Look sample |
| PD-L1 <50% | 1 year | **47.9%** | SHERLOC Model |
|  |  | **47.9%** | Alternate Model |
|  |  | 44.5% | Glans-Look sample |
|  | 3 year | **15.0%** | SHERLOC Model |
|  |  | **15.0%** | Alternate Model |
|  |  | 12.9% | Glans-Look sample |
|  | 5 year | **6.3%** | SHERLOC Model |
|  |  | **6.3%** | Alternate Model |
|  |  | 8.1% | Glans-Look sample |
| **External** | | | |
| total | 1 year | **74.3%** | SHERLOC Model |
|  |  | **74.1%** | Alternate Model |
|  |  | 65.3% | 57 |
|  |  | 64.6% | 55 |
|  |  | 64.6% | 58 |
|  | 3 years | **45.4%** | SHERLOC Model |
|  |  | **45.8%** | Alternate Model |
|  |  | 46.4% | 57 |
|  |  | 44.0% | 55 |
|  |  | 43.4% | 58 |
|  | 5 years | **30.3%** | SHERLOC Model |
|  |  | **30.7%** | Alternate Model |
|  |  | 37.6% | 57 |
|  |  | 34.4% | 56 |
|  |  | 36.0% | 55 |
|  |  | 34.7% | 58 |
|  |  | 30.4% | 59 |
| stage I | 1 year | **89.7%** | SHERLOC Model |
|  |  | **89.6%** | Alternate Model |
|  |  | 92.0% | 58 |
|  |  | 91.0% | 60 |
|  | 3 years | **67.4%** | SHERLOC Model |
|  |  | **67.2%** | Alternate Model |
|  |  | 77.9% | 58 |
|  |  | 72.0% | 60 |
|  | 5 years | **48.9%** | SHERLOC Model |
|  |  | **48.9%** | Alternate Model |
|  |  | 68.2% | 58 |
|  |  | 58.0% | 60 |
| stage II | 1 year | **92.6%** | SHERLOC Model |
|  |  | **94.5%** | Alternate Model |
|  |  | 80.3% | 58 |
|  |  | 79.0% | 60 |
|  | 3 years | **66.6%** | SHERLOC Model |
|  |  | **70.7%** | Alternate Model |
|  |  | 60.3% | 58 |
|  |  | 51.0% | 60 |
|  | 5 years | **47.5%** | SHERLOC Model |
|  |  | **49.4%** | Alternate Model |
|  |  | 47.5% | 58 |
|  |  | 40.0% | 60 |
| Stage III | 1 year | **87.0%** | SHERLOC Model |
|  |  | **86.5%** | Alternate Model |
|  |  | 76.9% | 58 |
|  |  | 63.0% | 60 |
|  |  | 90.0% | 61 |
|  | 3 years | **53.5%** | SHERLOC Model |
|  |  | **54.2%** | Alternate Model |
|  |  | 55.2% | 58 |
|  |  | 31.0% | 60 |
|  |  | 56.0% | 61 |
|  | 5 years | **36.3%** | SHERLOC Model |
|  |  | **36.6%** | Alternate Model |
|  |  | 41.4% | 58 |
|  |  | 21.0% | 60 |
| Stage IV | 1 year | **58.9%** | SHERLOC Model |
|  |  | **58.4%** | Alternate Model |
|  |  | 43.2% | 58 |
|  | 3 years | **27.5%** | SHERLOC Model |
|  |  | **27.7%** | Alternate Model |
|  |  | 21.7% | 58 |
|  | 5 years | **15.5%** | SHERLOC Model |
|  |  | **15.9%** | Alternate Model |
|  |  | 13.9% | 58 |
| EGFR | 1 year | **70.6%** | SHERLOC Model |
|  |  | **78.7%** | Alternate Model |
|  |  | 79.0% | 66 |
|  |  | 76.4% | 63 |
|  |  | 68.8% | 74 |
|  | 2 years | **48.8%** | SHERLOC Model |
|  |  | **48.9%** | Alternate Model |
|  |  | 41.0% | 66 |
|  |  | 58.0% | 63 |
|  |  | 49.7% | 74 |
|  | 3 years | **33.6%** | SHERLOC Model |
|  |  | **34.4%** | Alternate Model |
|  |  | 41.0% | 66 |
|  |  | 35.0% | 74 |
|  | 5 years | **14.9%** | SHERLOC Model |
|  |  | **16.9%** | Alternate Model |
|  |  | 18.0% | 66 |
| ALK | 1 year | **93.8%** | SHERLOC Model |
|  |  | **92.7%** | Alternate Model |
|  |  | 87.0% | 6 |
|  |  | 84.3% | 65 |
|  | 2 years | **80.8%** | SHERLOC Model |
|  |  | **81.3%** | Alternate Model |
|  |  | 73.0% | 6 |
|  |  | 72.5% | 65 |
|  | 3 years | **68.9%** | SHERLOC Model |
|  |  | **69.4%** | Alternate Model |
|  |  | 67.0% | 64 |
|  |  | 62.0% | 65 |
|  | 5 years | **51.2%** | SHERLOC Model |
|  |  | **51.8%** | Alternate Model |
|  |  | 59.5% | 64 |
|  |  | 45.0% | 65 |
| PD-L1 ≥50% | 1 year | **61.4%** | SHERLOC Model |
|  |  | **61.4%** | Alternate Model |
|  |  | 61.4% | 67 |
|  |  | 59.1% | 67 |
|  |  | 59.8% | 67 |
|  | 2 years | **44.1%** | SHERLOC Model |
|  |  | **44.1%** | Alternate Model |
|  |  | 44.7% | 67 |
|  |  | 45.7% | 67 |
|  | 3 years | **33.6%** | SHERLOC Model |
|  |  | **33.6%** | Alternate Model |
|  |  | 35.9% | 67 |
|  |  | 36.2% | 67 |
|  | 5 years | **22.6%** | SHERLOC Model |
|  |  | **22.6%** | Alternate Model |
|  |  | 25.9% | 67 |
|  |  | 26.9% | 68 |
| PD-L1 <50% | 1 year | **48.0%** | SHERLOC Model |
|  |  | **48.0%** | Alternate Model |
|  |  | 50.2% | 73 |
|  |  | 54.2% | 73 |
|  |  | 56.3% | 70 |
|  |  | 53.8% | 69 |
|  |  | 50.0% | 69 |
|  |  | 42.9% | 75 |
|  | 2 years | **26.4%** | SHERLOC Model |
|  |  | **26.4%** | Alternate Model |
|  |  | 34.7% | 73 |
|  |  | 33.0% | 73 |
|  |  | 36.4% | 69 |
|  |  | 31.5% | 69 |
|  | 3 years | **16.1%** | SHERLOC Model |
|  |  | **16.1%** | Alternate Model |
|  |  | 25.3% | 71 |
|  |  | 25.5% | 72 |
|  |  | 17.6% | 75 |
|  | 5 years | **7.1%** | SHERLOC Model |
|  |  | **7.1%** | Alternate Model |
|  |  | 13.4% | 71 |
|  |  | 14.4% | 72 |

^a^= Adjustment made for SCLC from SEER data^53^ of 1 year=1.15 3 years=1.16, 5 years=1.13

^b^= Weight of Local and Regional for SEER of 22.65% 77.35% from reference 57

^c^= Weight of PD-L1 1%-49% and PD-L1 <1% from reference 24, with 37.67% and 62.33% for survival estimates from 68, 56 and 2 year timepoints from 68 extracted with IPDfromKM

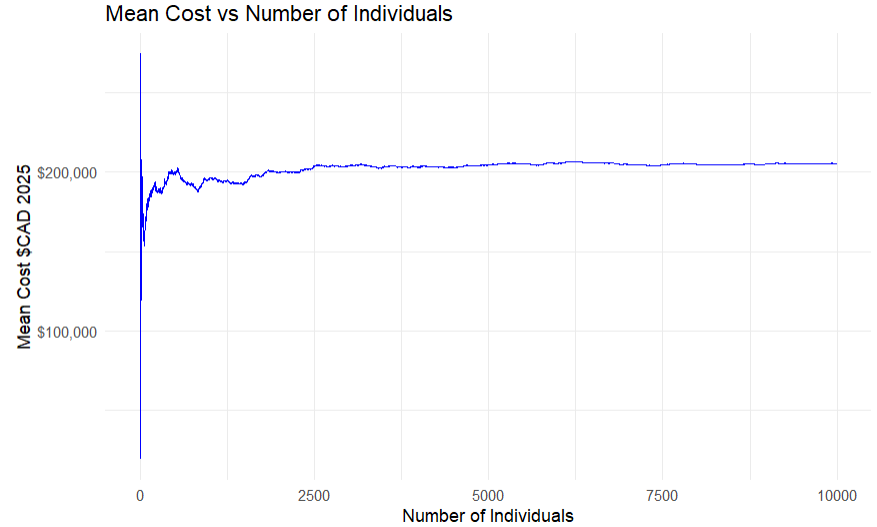

**Supplementary Figure 1:** First-order convergence plot, mean Cost per case vs number of simulated individuals.

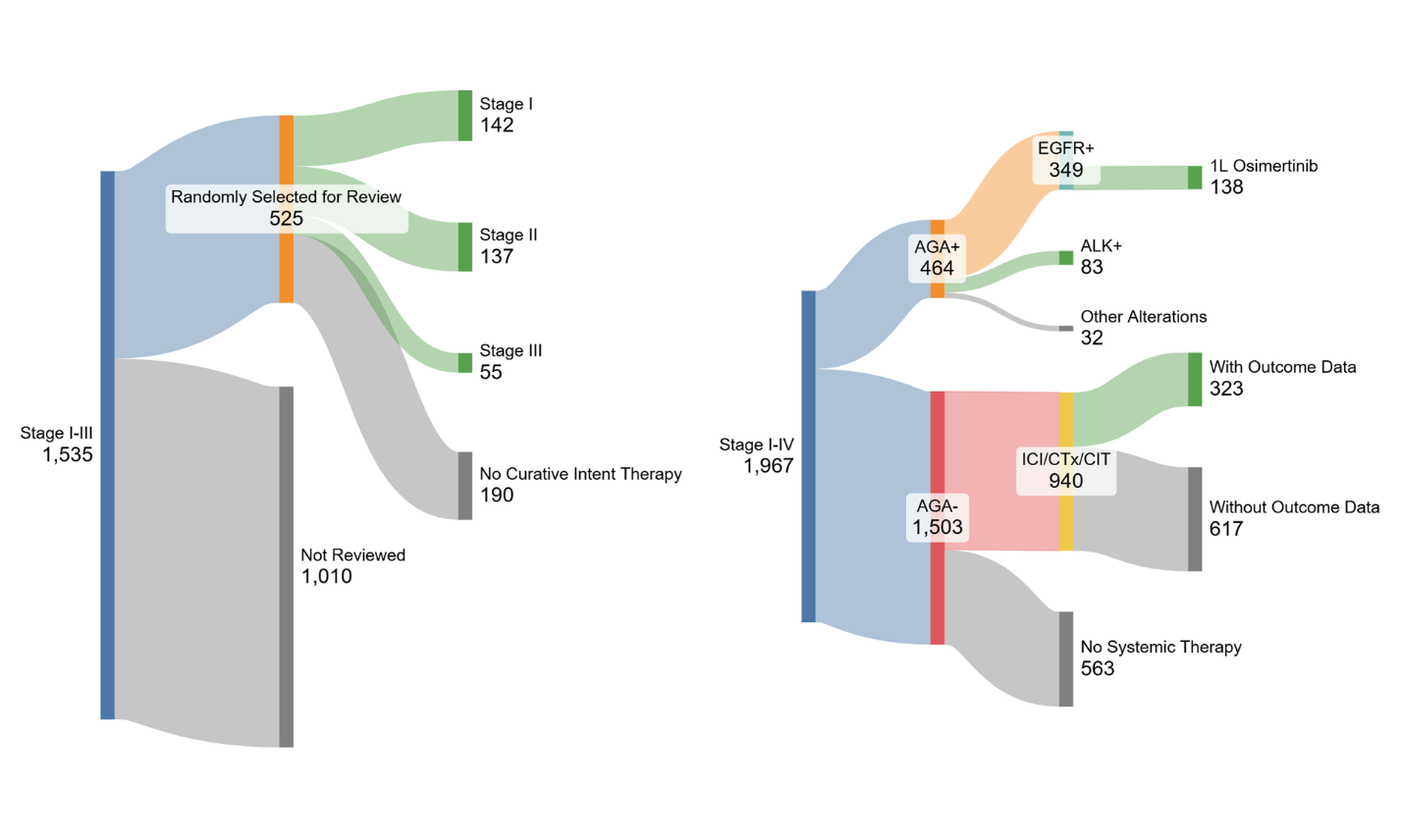

**Supplementary Figure 2:** Sankey plot of patient chart review inclusion exclusion

**Stage I-Death:**

| Distribution | AIC | RANK | BIC | Rank |
| --- | --- | --- | --- | --- |
| weibull | 718.5316 | 3 | 724.4291 | 3 |
| llogis | 719.0676 | 5 | 724.9651 | 5 |
| lognormal | 725.1689 | 7 | 731.0664 | 7 |
| exp | **718.1316** | **1** | **721.0804** | **1** |
| gamma | 718.7068 | 4 | 724.6043 | 4 |
| gengamma | 720.3709 | 6 | 729.2172 | 6 |
| gompertz | 718.479 | 2 | 724.3766 | 2 |

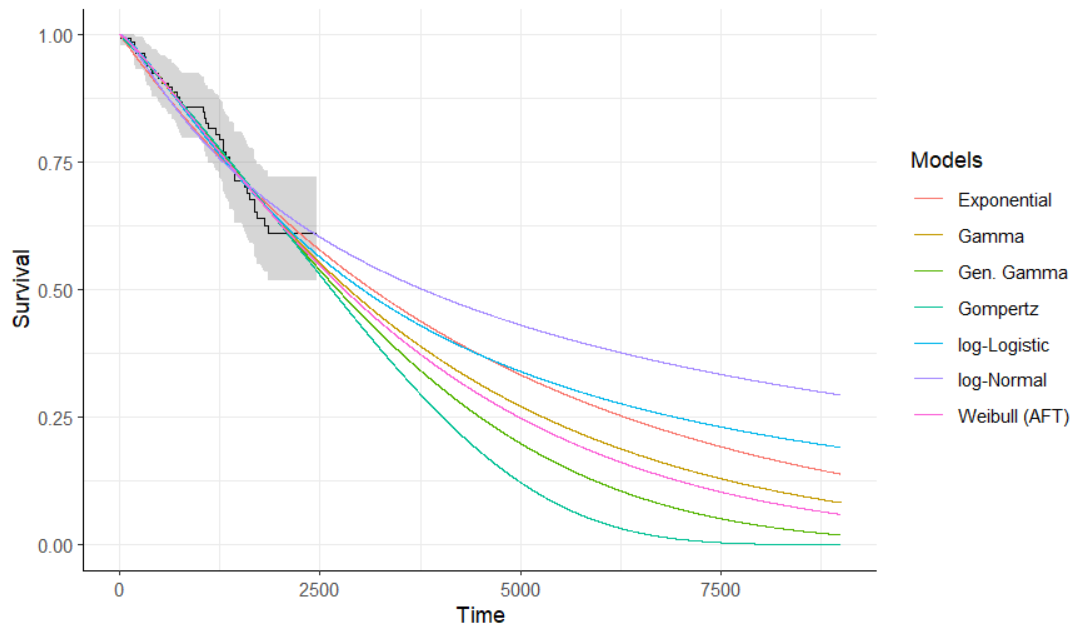

**Supplementary Figure 3:**

**Stage I-Stage IV:**

| Distribution | AIC | Rank | BIC | Rank |
| --- | --- | --- | --- | --- |
| weibull | 1031.681647 | 6 | 1040.527927 | 6 |
| llogis | 1028.44841 | 3 | 1037.29469 | 3 |
| lognormal | **1025.46229** | **1** | **1034.30857** | **1** |
| exp | 1029.723829 | 4 | 1035.621349 | 2 |
| gamma | 1031.722605 | 7 | 1040.568885 | 7 |
| gengamma | 1026.199897 | 2 | 1037.994937 | 4 |
| gompertz | 1029.934664 | 5 | 1038.780943 | 5 |

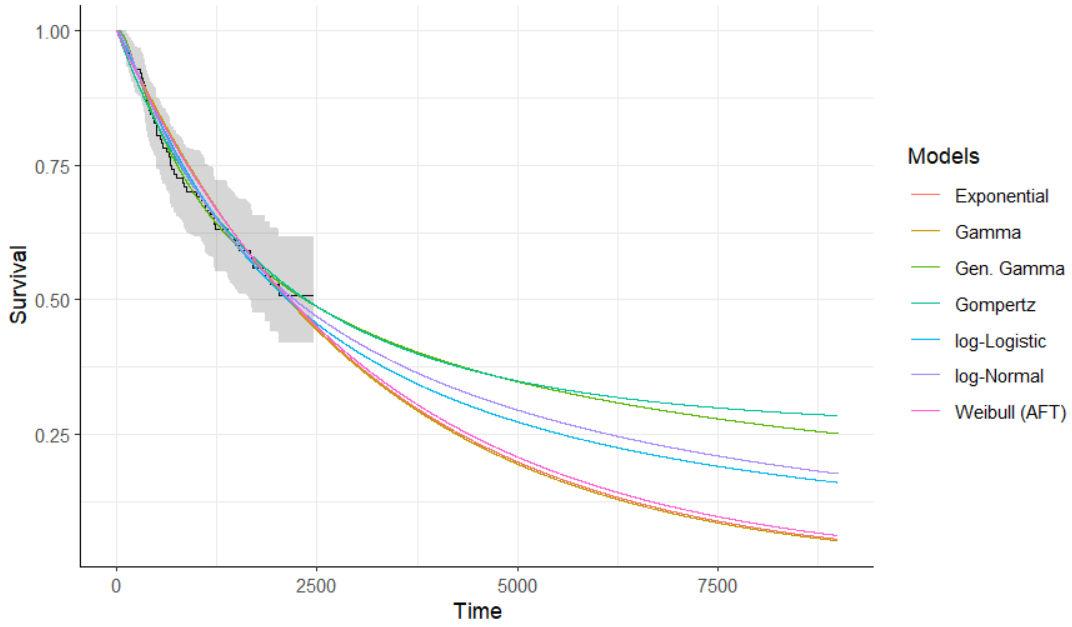

**Supplementary Figure 4:**

**Stage II-Death:**

| Distribution | AIC | Rank | BIC | Rank |
| --- | --- | --- | --- | --- |
| weibull | 417.0584 | 3 | 422.8541 | 3 |
| llogis | 418.1017 | 5 | 423.8974 | 5 |
| lognormal | 424.1081 | 7 | 429.9038 | 7 |
| exp | **416.4759** | **2** | **419.3737** | **1** |
| gamma | 417.485 | 4 | 423.2807 | 4 |
| gengamma | 418.2172 | 6 | 426.9108 | 6 |
| gompertz | 414.7278 | 1 | 420.5235 | 2 |

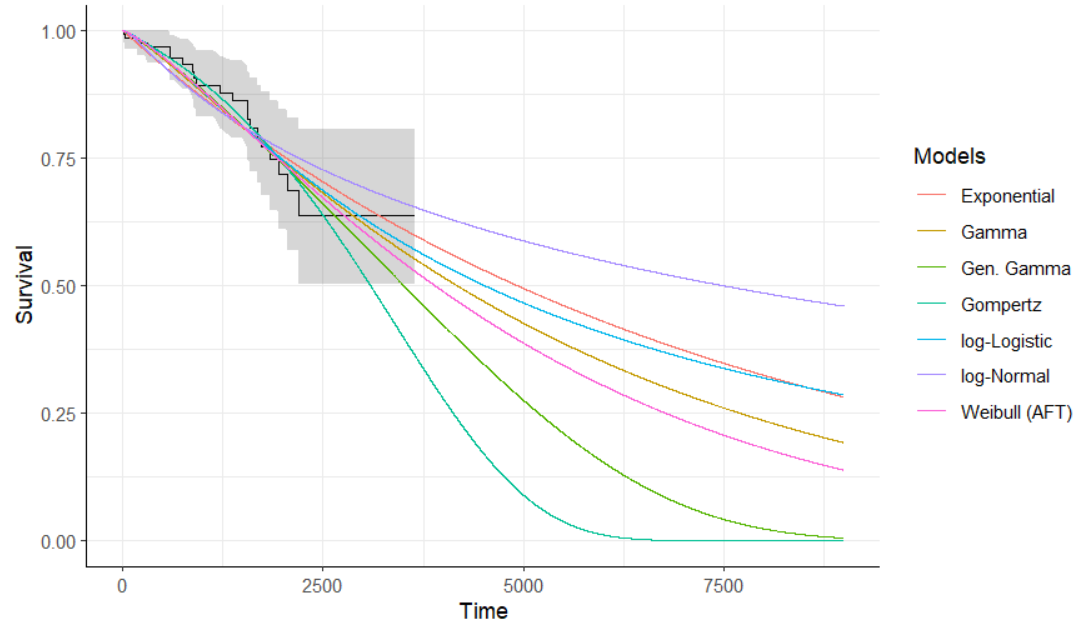

**Supplementary Figure 5:**

**Stage II-Stage IV:**

| Distribution | AIC | Rank | BIC | Rank |
| --- | --- | --- | --- | --- |
| weibull | 1291.923 | 6 | 1303.484 | 6 |
| llogis | 1285.504 | 4 | 1297.065 | 4 |
| lognormal | 1281.301 | 2 | 1292.862 | 1 |
| exp | 1291.34 | 5 | 1300.011 | 5 |
| gamma | 1292.772 | 7 | 1304.333 | 7 |
| gengamma | **1279.715** | **1** | **1294.167** | **2** |
| gompertz | 1284.619 | 3 | 1296.181 | 3 |

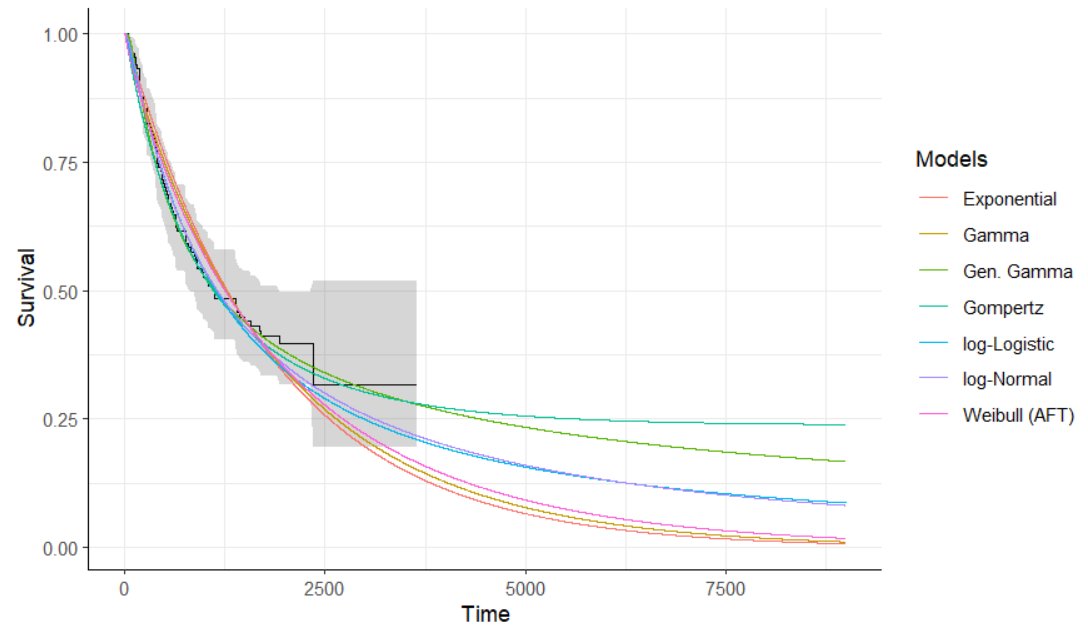

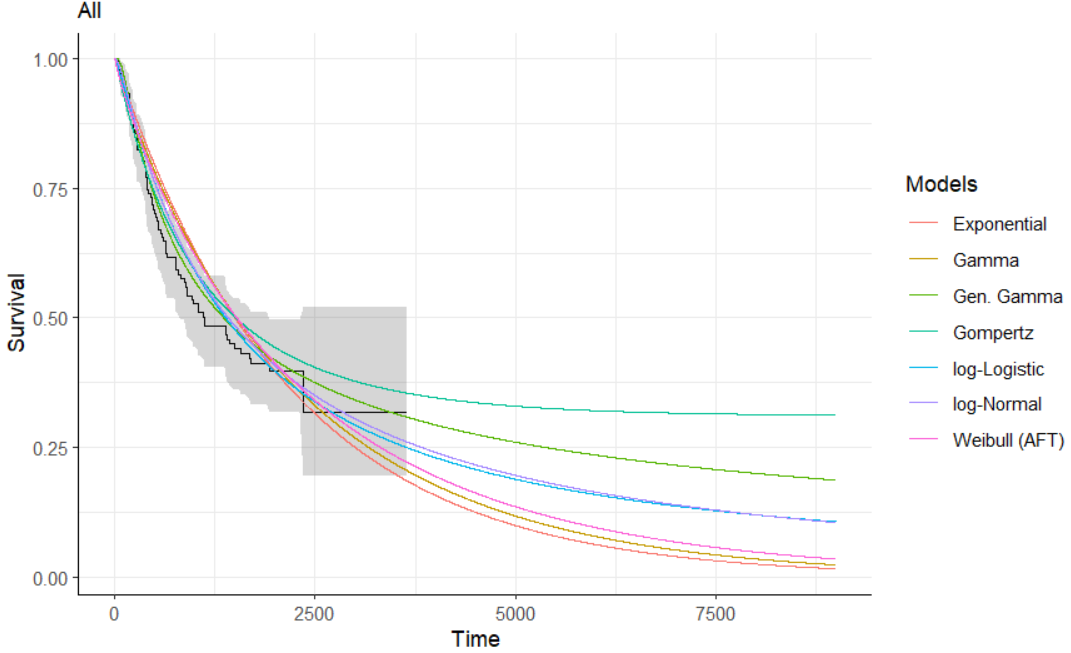

**Supplementary Figure 6:**

**Stage III-Death:**

| Distribution | AIC | Rank | BIC | Rank |
| --- | --- | --- | --- | --- |
| weibull | 191.3541 | 5 | 195.2947 | 5 |
| llogis | 191.4813 | 6 | 195.4219 | 6 |
| lognormal | **190.9613** | **2** | **194.9019** | **2** |
| exp | 189.4188 | 1 | 191.3891 | 1 |
| gamma | 191.3462 | 4 | 195.2868 | 4 |
| gengamma | 192.8967 | 7 | 198.8076 | 7 |
| gompertz | 191.3055 | 3 | 195.2461 | 3 |

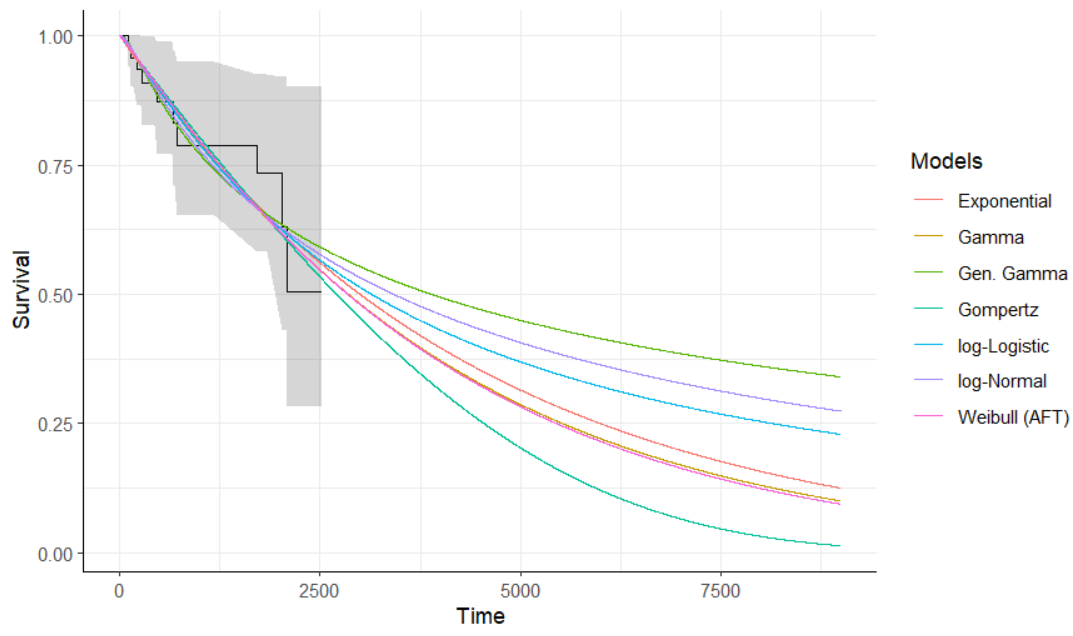

**Supplementary Figure 7:**

**Stage III-Stage IV:**

| Distribution | AIC | Rank | BIC | Rank |
| --- | --- | --- | --- | --- |
| weibull | 539.4327 | 5 | 543.3733 | 5 |
| llogis | 534.1826 | 4 | 538.1232 | 4 |
| lognormal | 532.4874 | 3 | 536.428 | 2 |
| exp | 541.6832 | 7 | 543.6535 | 6 |
| gamma | 540.9713 | 6 | 544.9119 | 7 |
| gengamma | **531.3185** | **1** | **537.2294** | **3** |
| gompertz | 532.389 | 2 | 536.3296 | 1 |

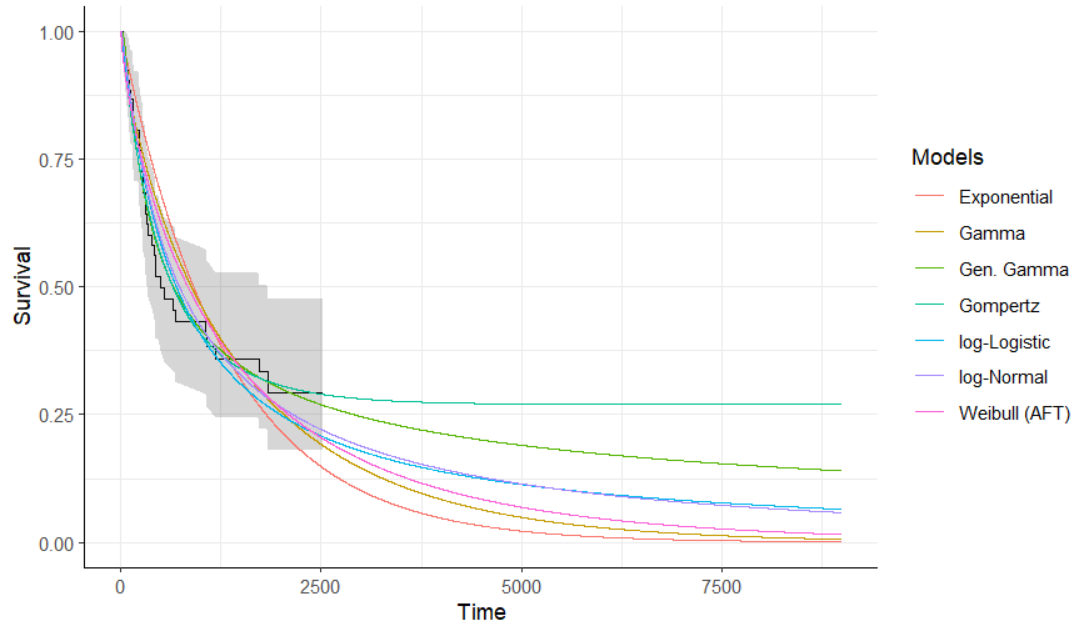

**Supplementary Figure 8:**

**EGFR 1L-BSC:**

| Distribution | AIC | Rank | BIC | Rank |
| --- | --- | --- | --- | --- |
| weibull | 1054.185 | 6 | 1062.788 | 5 |
| llogis | 1050.58 | 1 | 1059.182 | 2 |
| lognormal | 1051.939 | 2 | 1060.541 | 3 |
| exp | **1052.228** | **3** | **1057.963** | **1** |
| gamma | 1054.228 | 7 | 1062.831 | 6 |
| gengamma | 1053.52 | 4 | 1064.99 | 7 |
| gompertz | 1053.608 | 5 | 1062.211 | 4 |

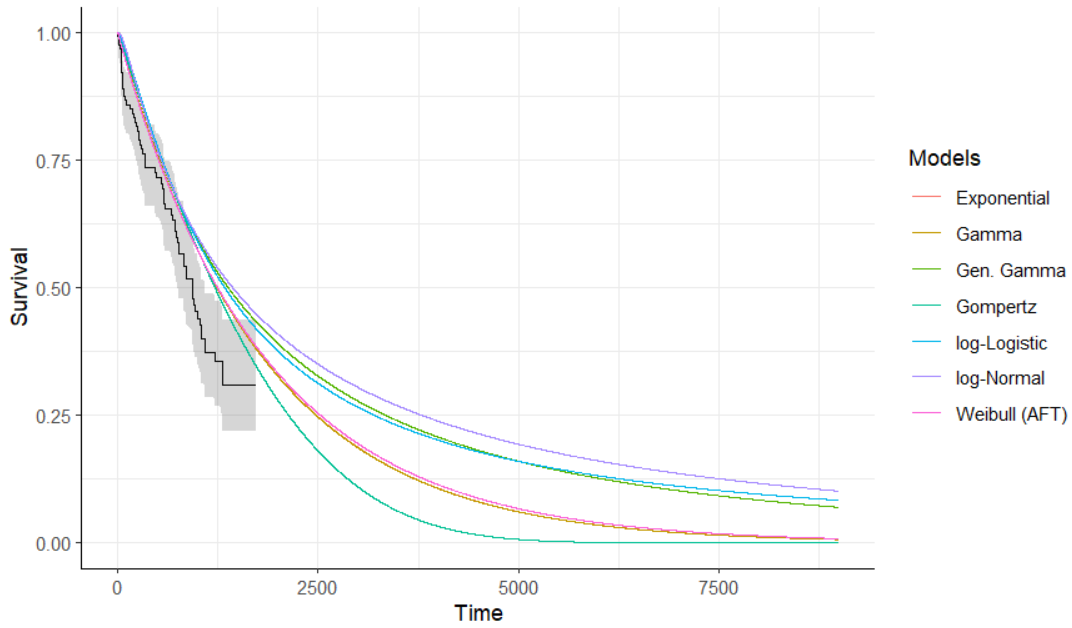

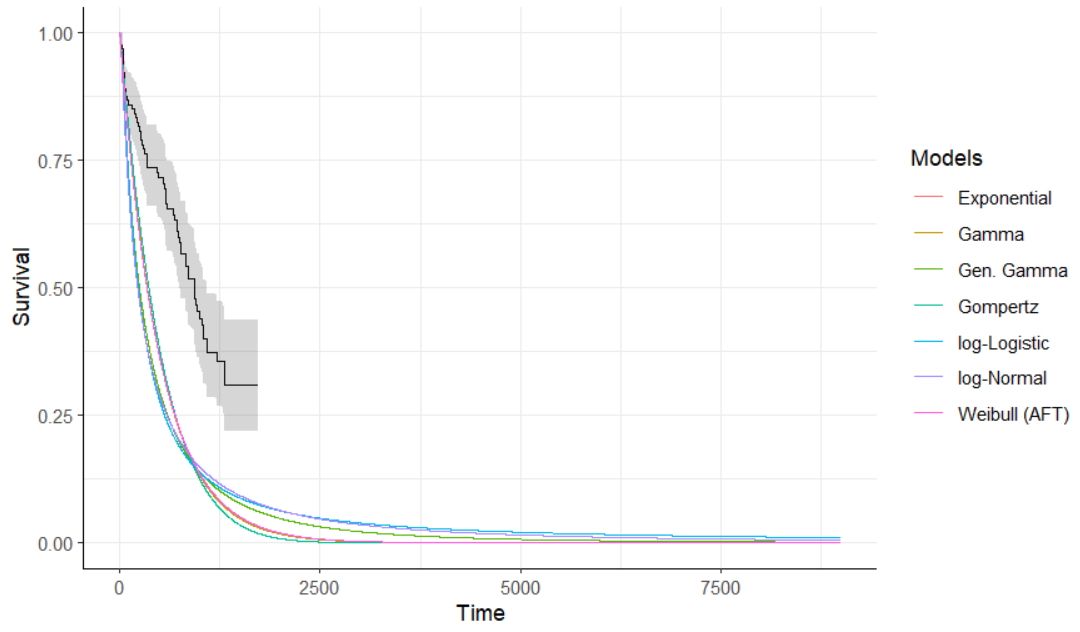

**Supplementary Figure 9:**

**EGFR 1L-2L:**

| Distribution | AIC | Rank | BIC | Rank |
| --- | --- | --- | --- | --- |
| weibull | 678.5262465 | 5 | 687.1288499 | 5 |
| llogis | 678.1902192 | 3 | 686.7928226 | 3 |
| lognormal | 678.1468218 | 2 | 686.7494251 | 2 |
| exp | **676.5309914** | **1** | **682.2660603** | **1** |
| gamma | 678.5309862 | 6 | 687.1335895 | 6 |
| gengamma | 679.8750495 | 7 | 691.3451873 | 7 |
| gompertz | 678.301154 | 4 | 686.9037573 | 4 |

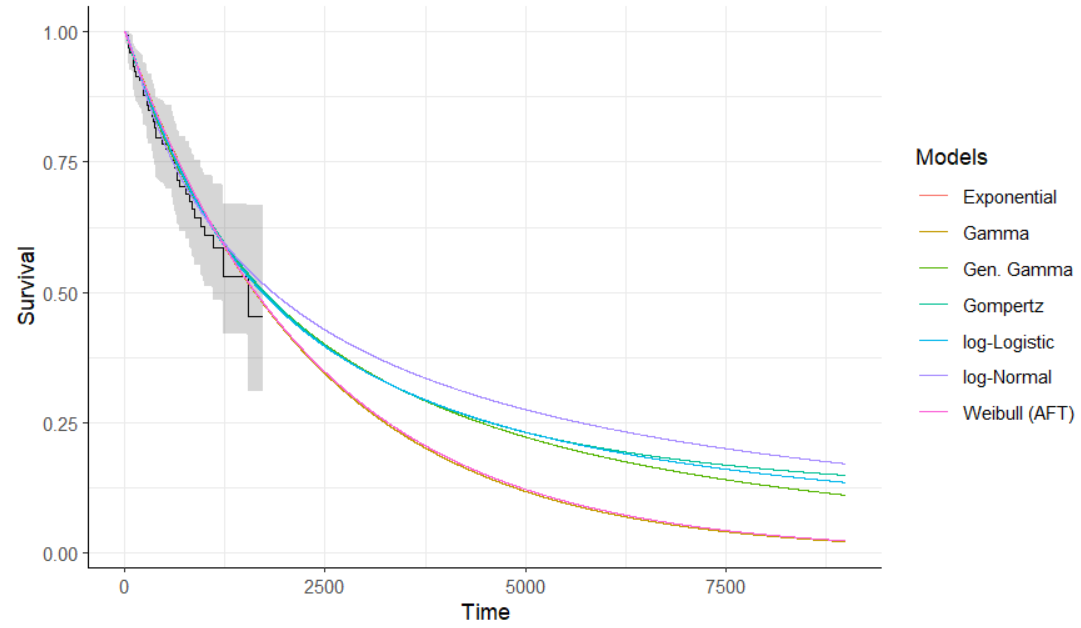

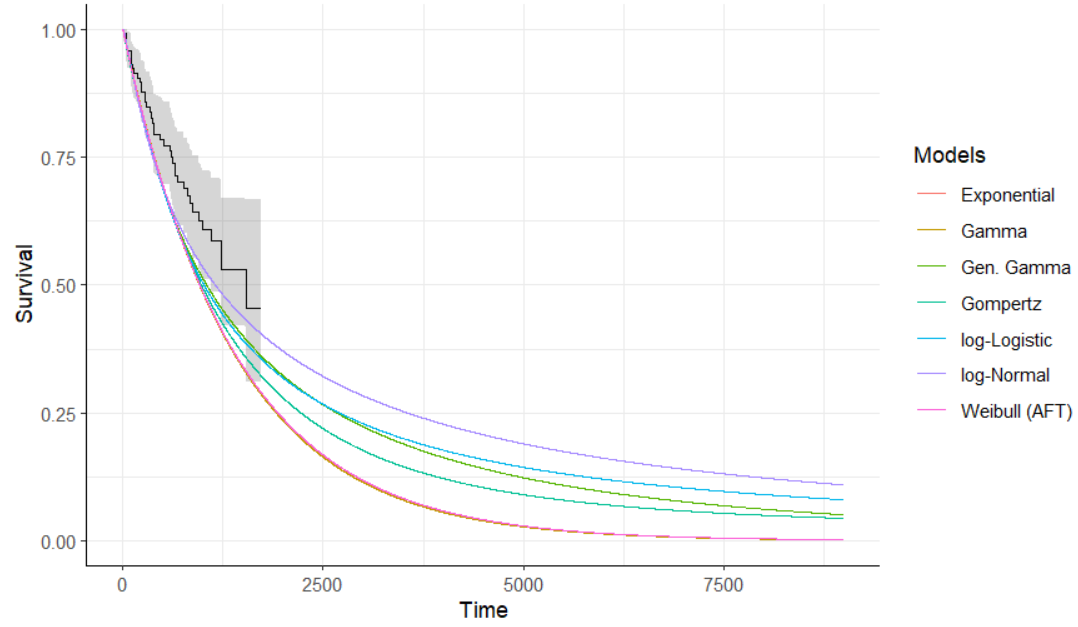

**Supplementary Figure 10:**

**ALK 1L-BSC:**

| Distribution | AIC | Rank | BIC | Rank |
| --- | --- | --- | --- | --- |
| weibull | 240.8705 | 3 | 243.3867 | 3 |
| llogis | 242.1582 | 5 | 244.6744 | 5 |
| lognormal | 244.7572 | 7 | 247.2734 | 7 |
| exp | **240.2164** | **1** | **241.4744** | **1** |
| gamma | 240.6379 | 2 | 243.1541 | 2 |
| gengamma | 242.4997 | 6 | 246.274 | 6 |
| gompertz | 241.7821 | 4 | 244.2983 | 4 |

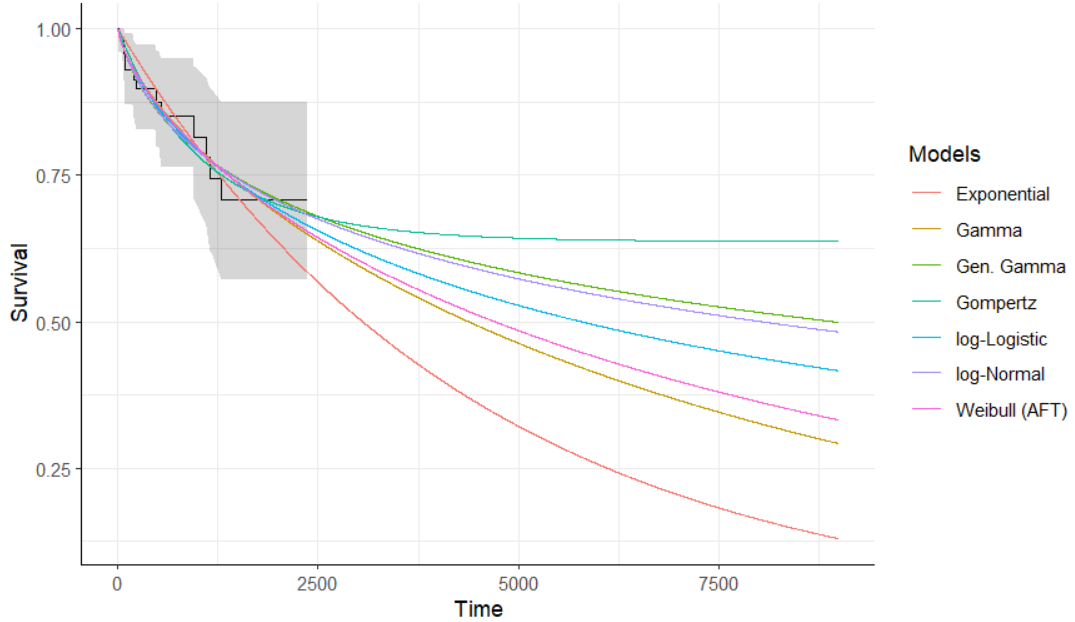

**Supplementary Figure 11:**

**ALK 1L-2L:**

| Distribution | AIC | Rank | BIC | Rank |
| --- | --- | --- | --- | --- |
| weibull | 456.2843 | 5 | 460.8376 | 5 |
| llogis | 455.8134 | 3 | 460.3667 | 3 |
| lognormal | **454.9725** | **2** | **459.5258** | **2** |
| exp | 454.2984 | 1 | 456.5751 | 1 |
| gamma | 456.2981 | 6 | 460.8514 | 6 |
| gengamma | 456.9684 | 7 | 463.7984 | 7 |
| gompertz | 456.1356 | 4 | 460.6889 | 4 |

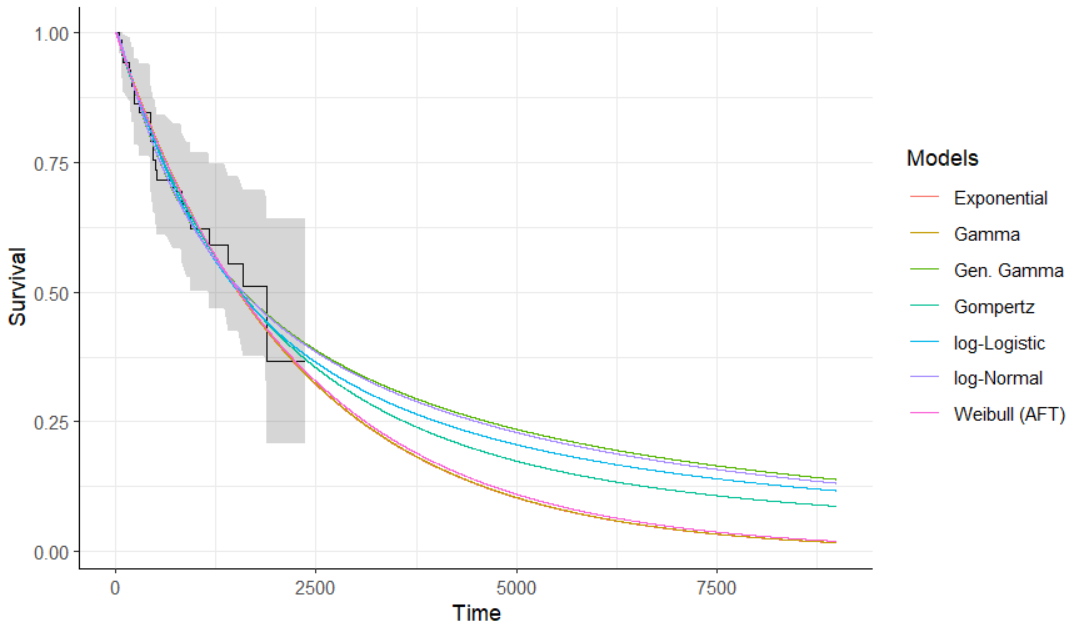

**Supplementary Figure 12:**

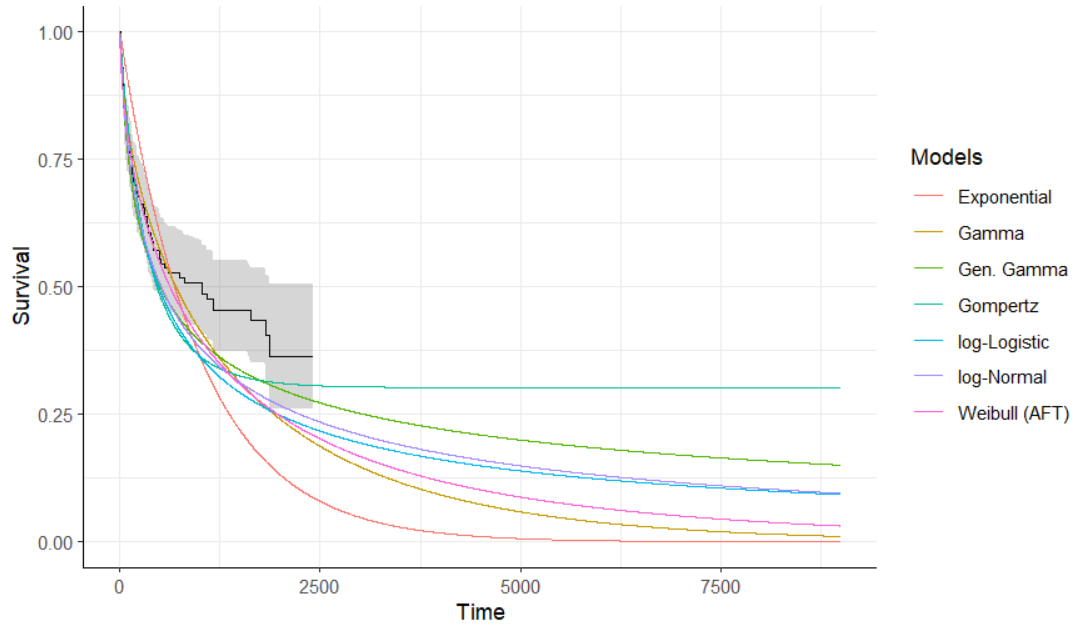

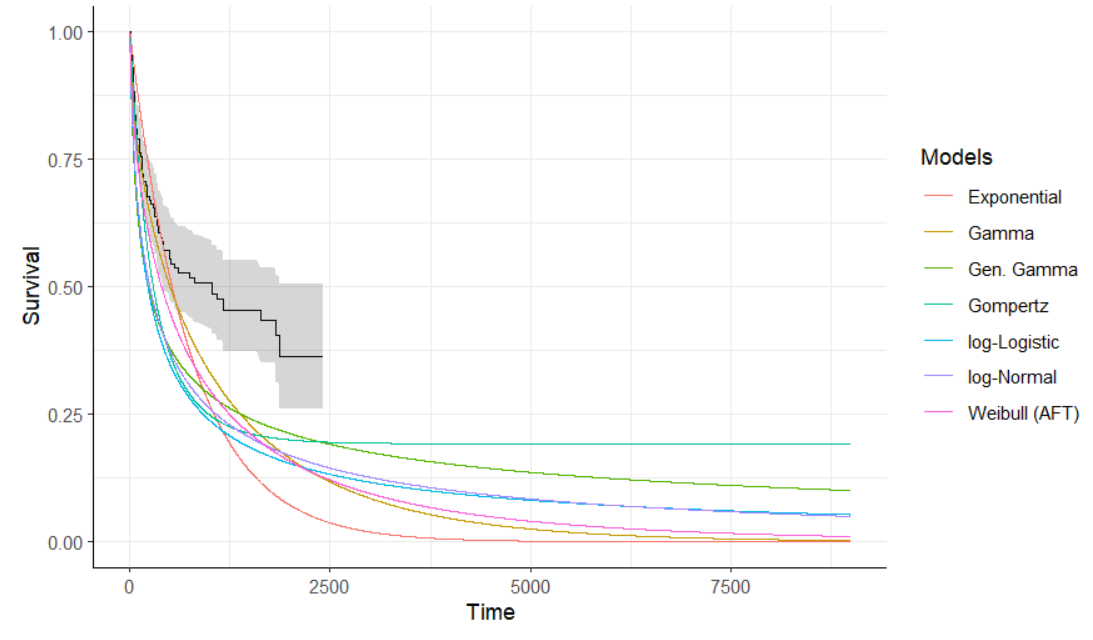

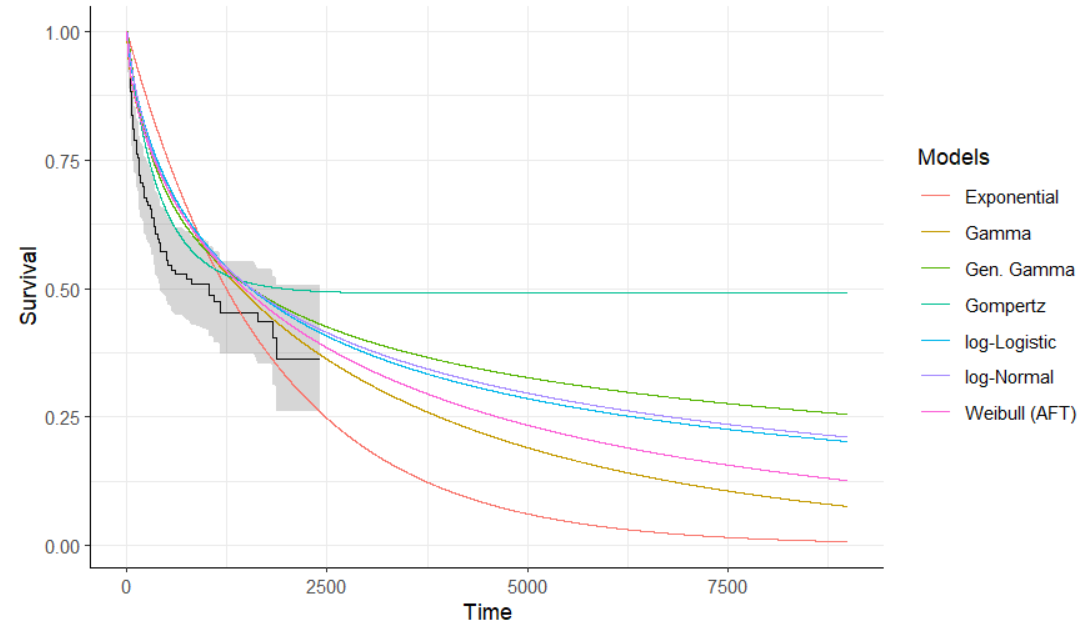

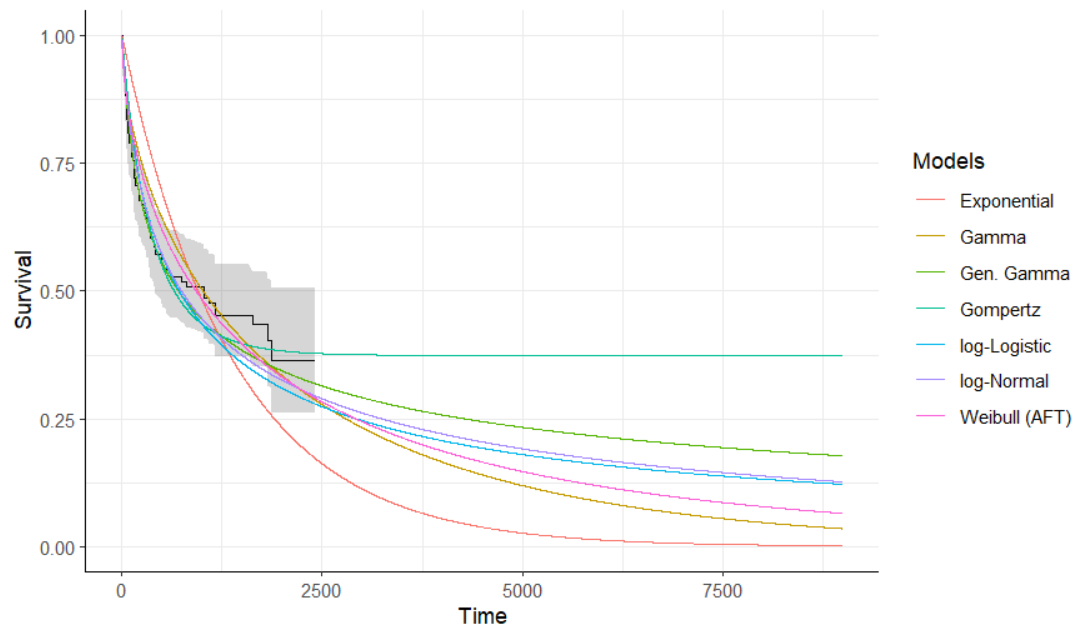
**PD-L1** ≥**50% 1L-BSC:**

| Distribution | AIC | Rank | BIC | Rank |
| --- | --- | --- | --- | --- |
| weibull | 1195.47715 | 5 | 1210.661913 | 5 |
| llogis | 1188.727527 | 3 | 1203.91229 | 3 |
| lognormal | **1184.929644** | **1** | **1200.114407** | **1** |
| exp | 1227.924958 | 7 | 1240.072769 | 7 |
| gamma | 1199.820736 | 6 | 1215.005499 | 6 |
| gengamma | 1184.999375 | 2 | 1203.22109 | 2 |
| gompertz | 1190.685976 | 4 | 1205.870739 | 4 |

**Supplementary Figure 13:**

**PD-L1** ≥**50% 1L-2L:**

| Distribution | AIC | Rank | BIC | Rank |
| --- | --- | --- | --- | --- |
| weibull | 758.3483 | 5 | 767.4786 | 6 |
| llogis | 755.3347 | 4 | 764.465 | 3 |
| lognormal | **752.5925** | **2** | **761.7228** | **2** |
| exp | 759.5048 | 7 | 765.5916 | 4 |
| gamma | 759.184 | 6 | 768.3143 | 7 |
| gengamma | 753.6382 | 3 | 765.8119 | 5 |
| gompertz | 752.3755 | 1 | 761.5057 | 1 |

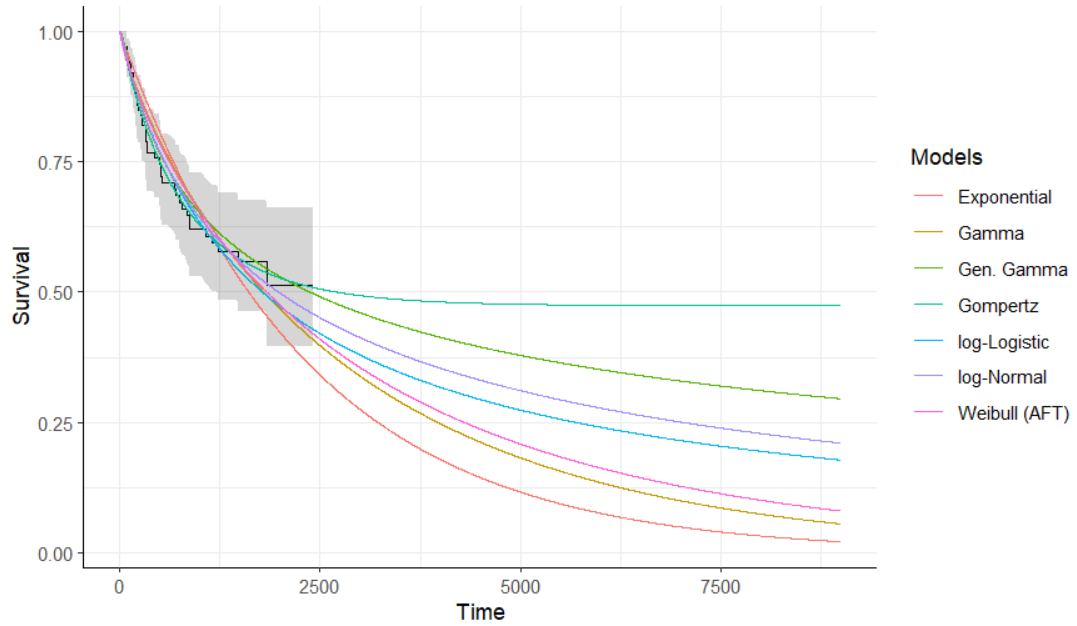

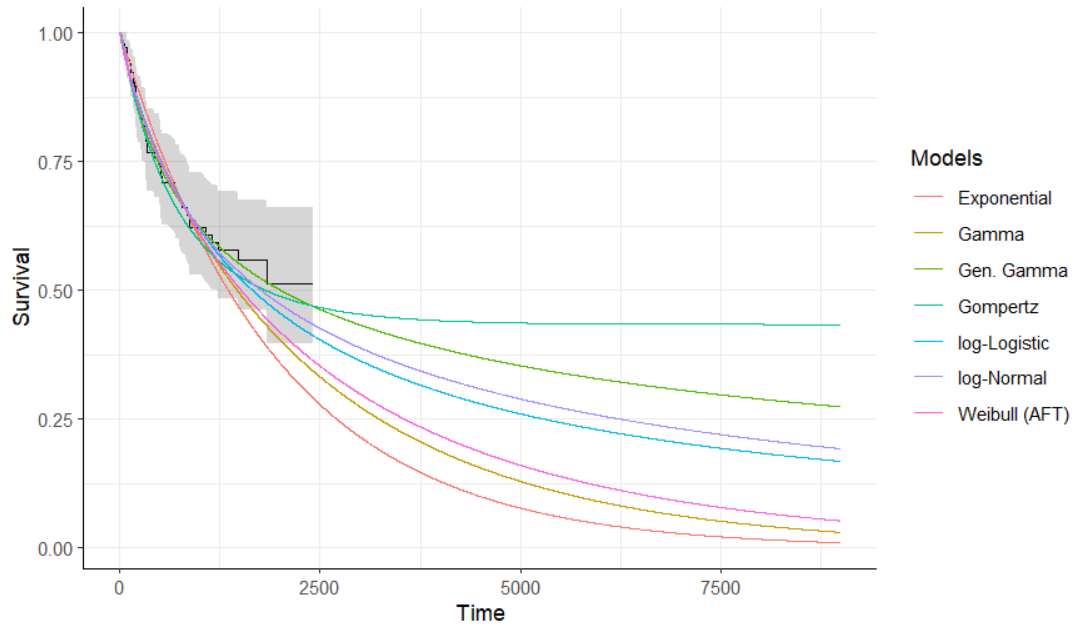

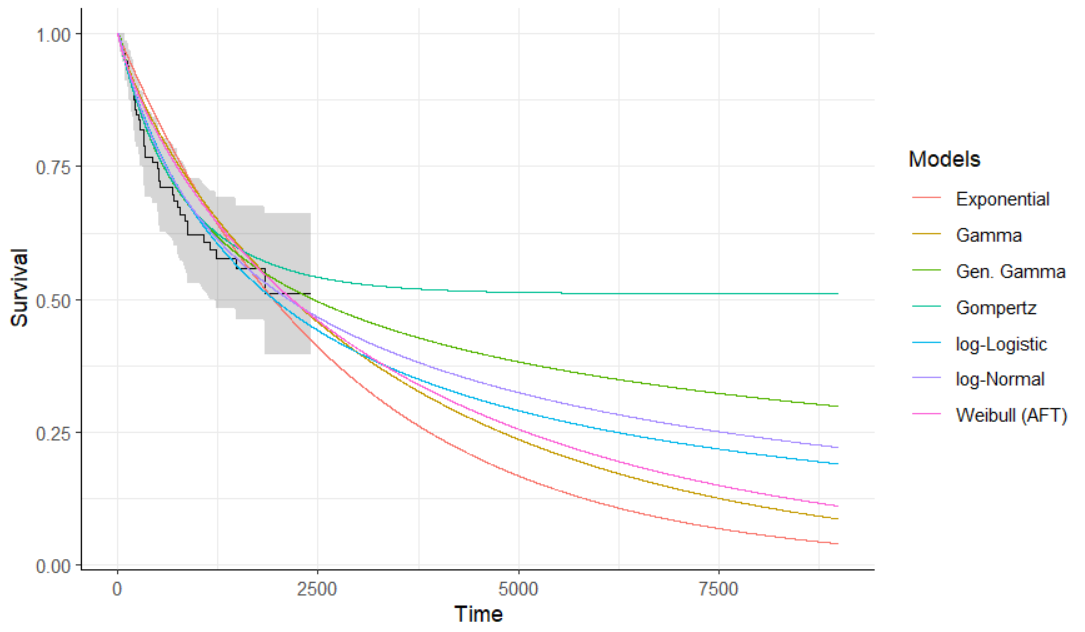

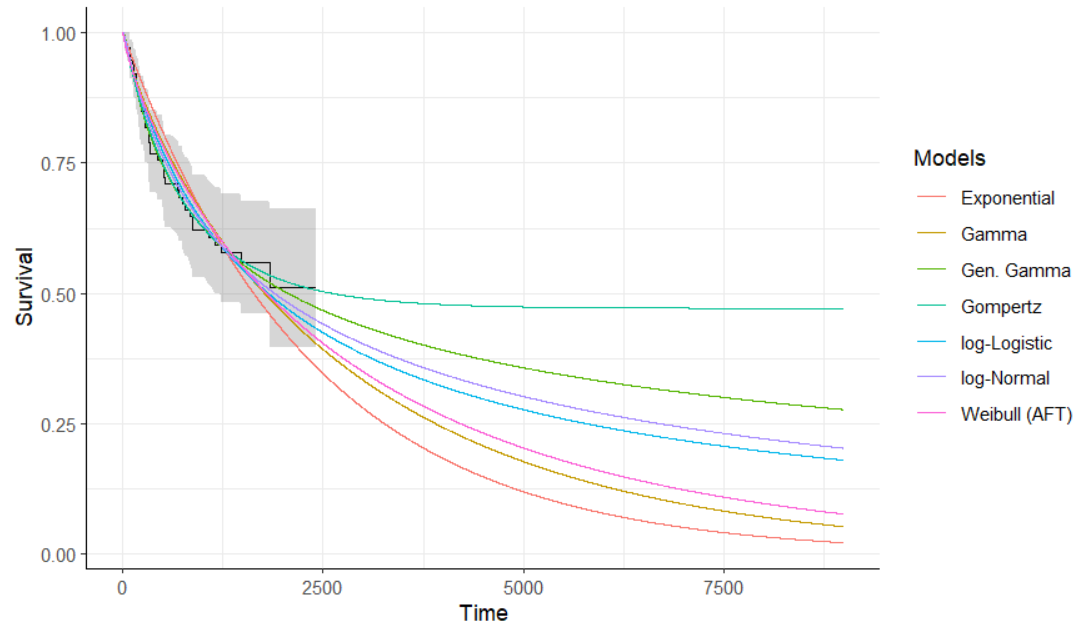

**Supplementary Figure 14:**

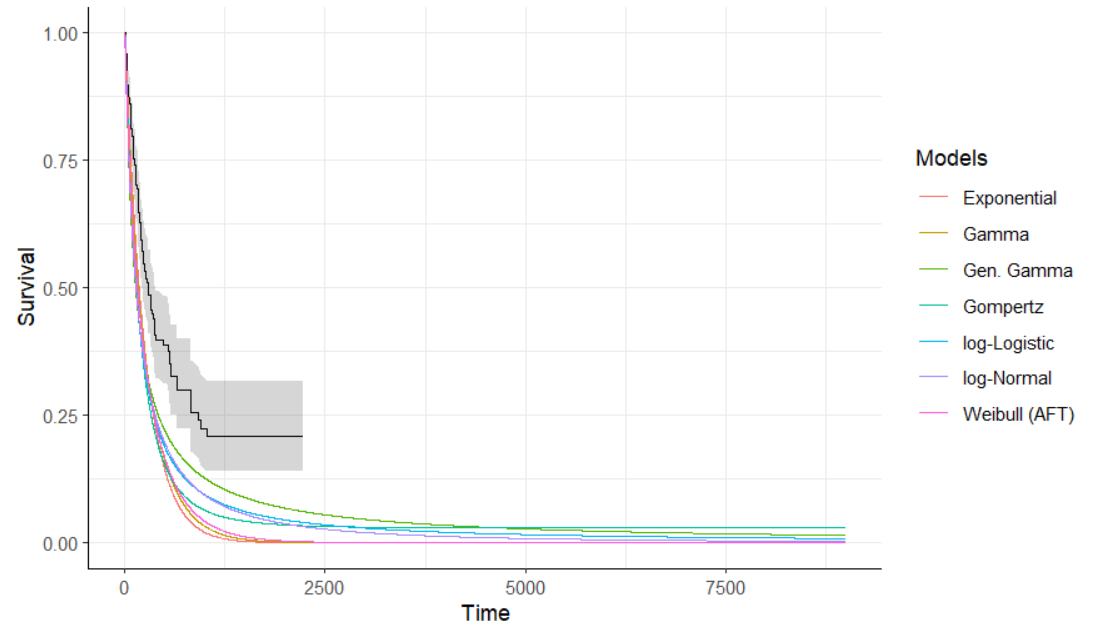

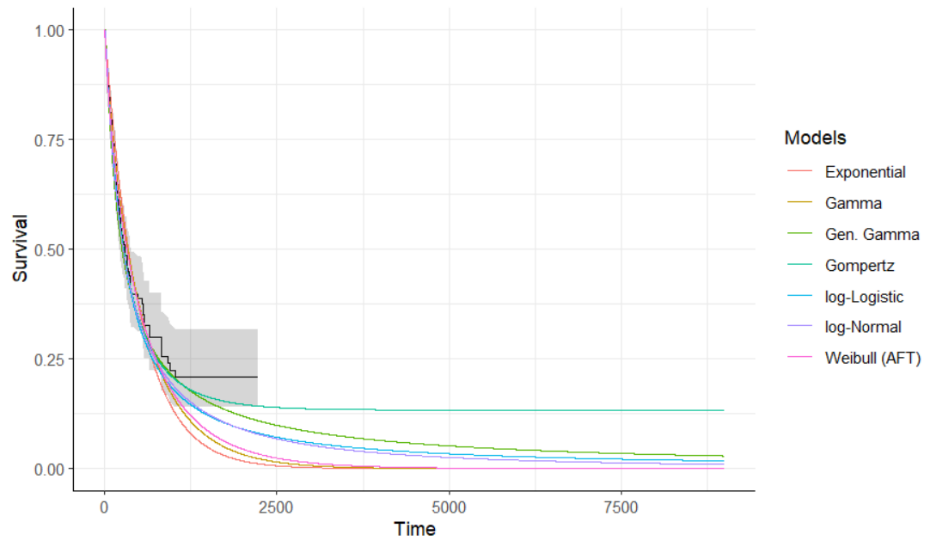

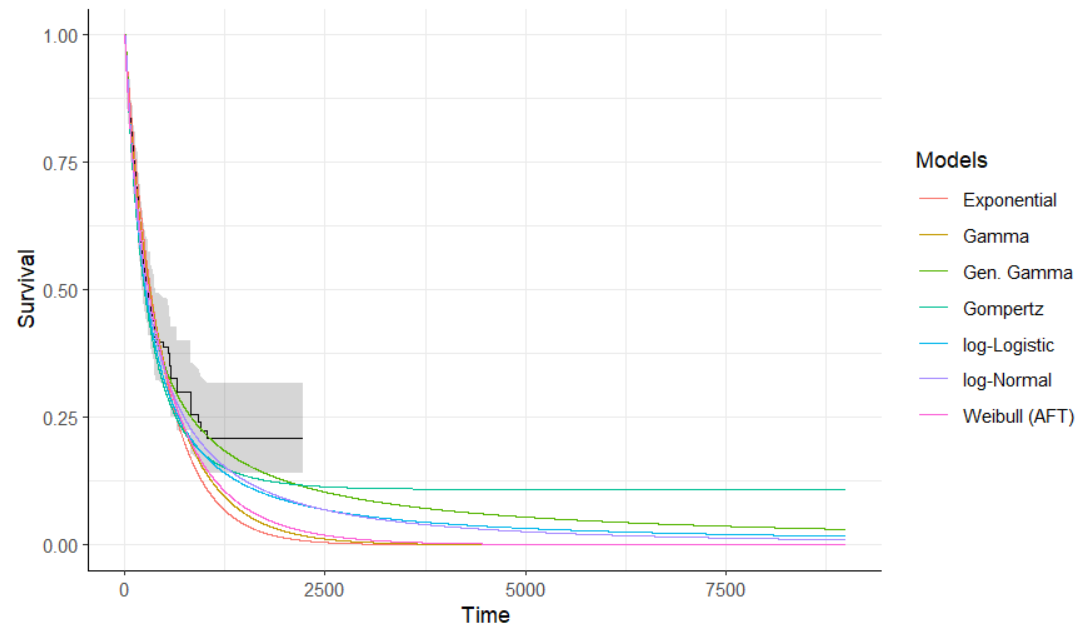

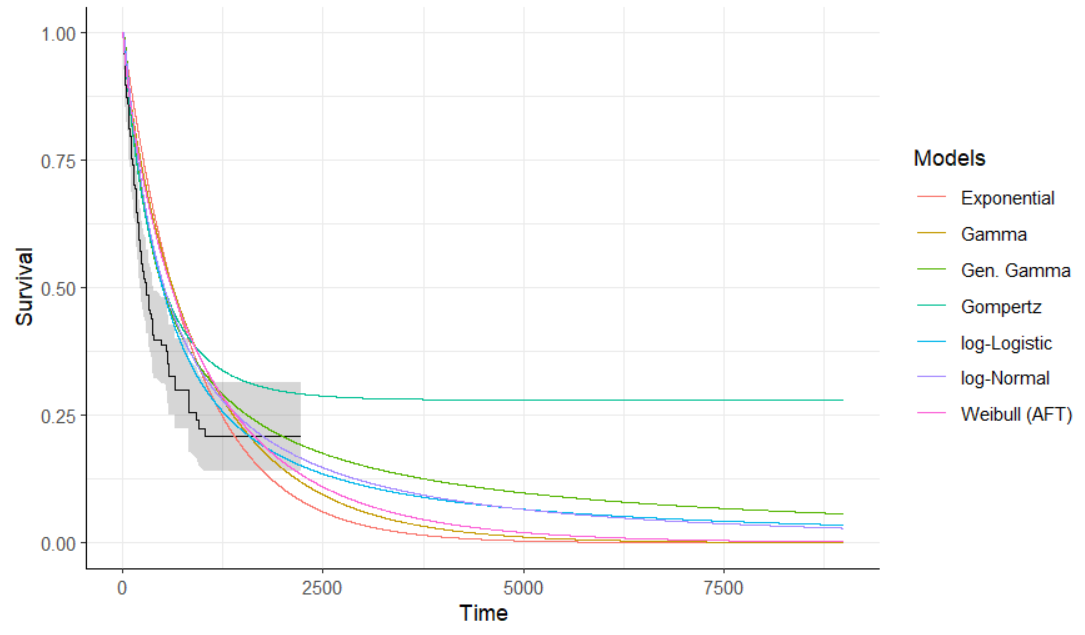
**PD-L1** <**50% 1L-BSC:**

| Distribution | AIC | Rank | BIC | Rank |
| --- | --- | --- | --- | --- |
| weibull | 1469.226 | 5 | 1484.846 | 5 |
| llogis | 1457.93 | 4 | 1473.55 | 3 |
| lognormal | **1455.222** | **1** | **1470.842** | **1** |
| exp | 1474.473 | 7 | 1486.969 | 6 |
| gamma | 1472.102 | 6 | 1487.722 | 7 |
| gengamma | 1455.779 | 3 | 1474.523 | 4 |
| gompertz | 1455.762 | 2 | 1471.382 | 2 |

**Supplementary Figure 15:**

**PD-L1** <**50% 1L-2L:**

| Distribution | AIC | Rank | BIC | Rank |
| --- | --- | --- | --- | --- |
| weibull | 908.5155237 | 6 | 917.8874156 | 6 |
| llogis | 904.7536114 | 2 | 914.1255034 | 3 |
| lognormal | **903.3978711** | **1** | **912.769763** | **1** |
| exp | 907.3201679 | 4 | 913.5680959 | 2 |
| gamma | 907.8349198 | 5 | 917.2068118 | 4 |
| gengamma | 905.3675004 | 3 | 917.8633564 | 5 |
| gompertz | 909.3197629 | 7 | 918.6916549 | 7 |

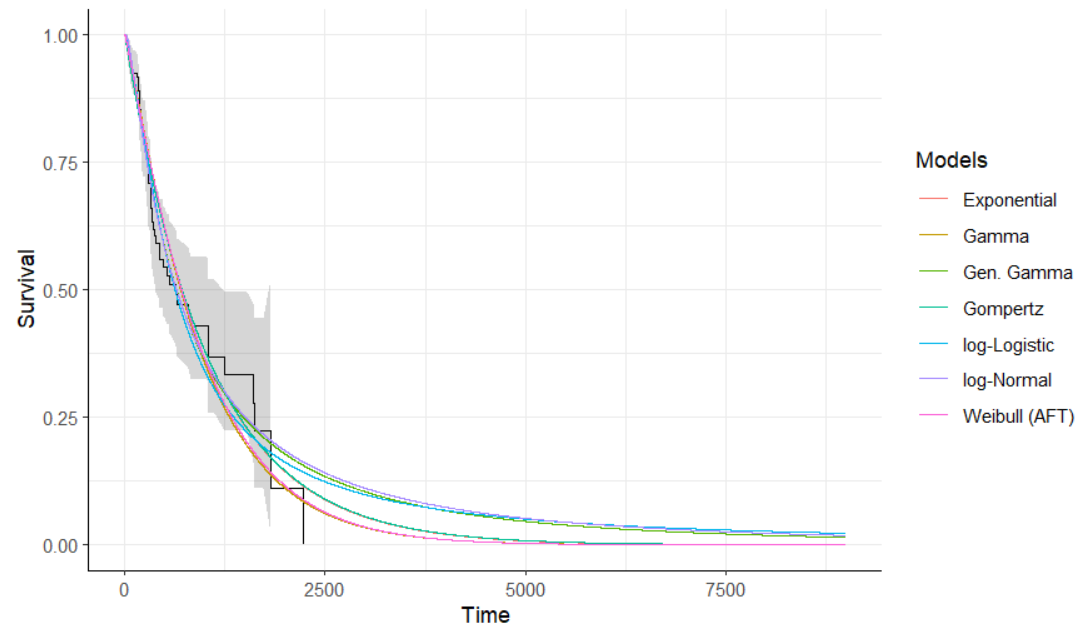

**Supplementary Figure 16:**

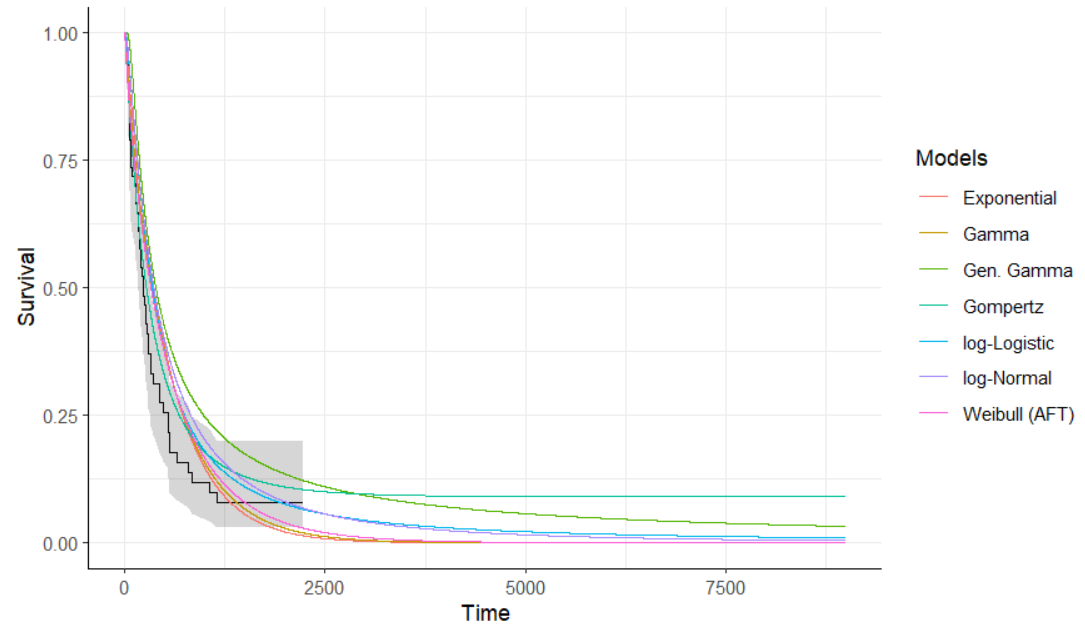

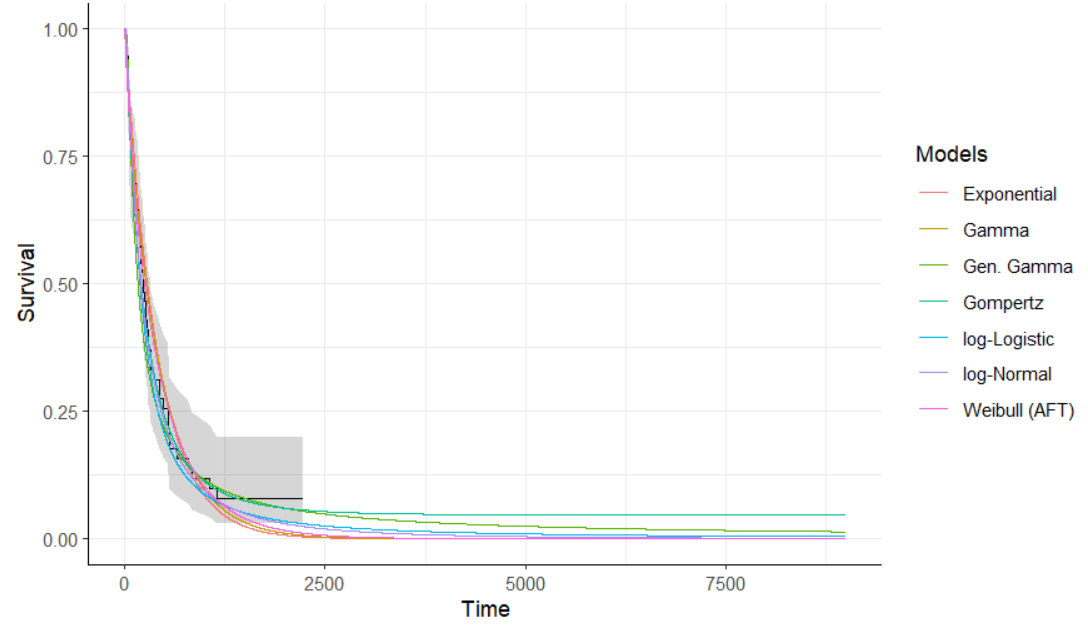
**PD-L1** <**50% 2L-BSC:**

| Distribution | AIC | Rank | BIC | Rank |
| --- | --- | --- | --- | --- |
| weibull | 711.5610945 | 5 | 719.7332995 | 6 |
| llogis | 699.8132186 | 3 | 707.9854236 | 2 |
| lognormal | **699.7427334** | **2** | **707.9149385** | **1** |
| exp | 711.9908791 | 6 | 718.1200329 | 5 |
| gamma | 713.1405141 | 7 | 721.3127192 | 7 |
| gengamma | 697.8413832 | 1 | 708.0566395 | 3 |
| gompertz | 703.7228324 | 4 | 711.8950375 | 4 |

**Supplementary Figure 17:**

**PD-L1** ≥**50% 2L-BSC:**

| Distribution | AIC | Rank | BIC | Rank |
| --- | --- | --- | --- | --- |
| weibull | 504.8916 | 6 | 508.414 | 6 |
| llogis | **496.0917** | **1** | **499.6141** | **1** |
| lognormal | 499.1004 | 2 | 502.6228 | 2 |
| exp | 503.0224 | 5 | 504.7836 | 4 |
| gamma | 504.9672 | 7 | 508.4896 | 7 |
| gengamma | 501.0961 | 4 | 506.3797 | 5 |
| gompertz | 499.77 | 3 | 503.2924 | 3 |

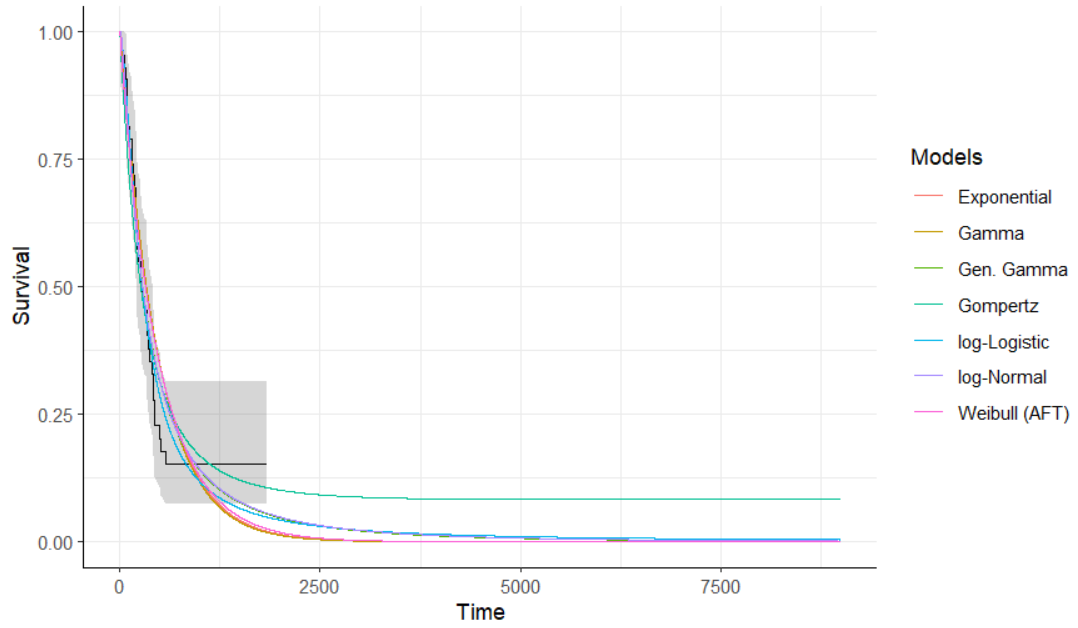

**Supplementary Figure 18:**

**EGFR 2L-BSC:**

| Distribution | AIC | Rank | BIC | Rank |
| --- | --- | --- | --- | --- |
| weibull | 484.1674 | 7 | 487.5452 | 6 |
| llogis | 481.6094 | 1 | 484.9872 | 2 |
| lognormal | 482.3518 | 3 | 485.7295 | 3 |
| exp | **482.1931** | **2** | **483.8819** | **1** |
| gamma | 483.9744 | 6 | 487.3521 | 5 |
| gengamma | 483.9055 | 5 | 488.9721 | 7 |
| gompertz | 483.7258 | 4 | 487.1036 | 4 |

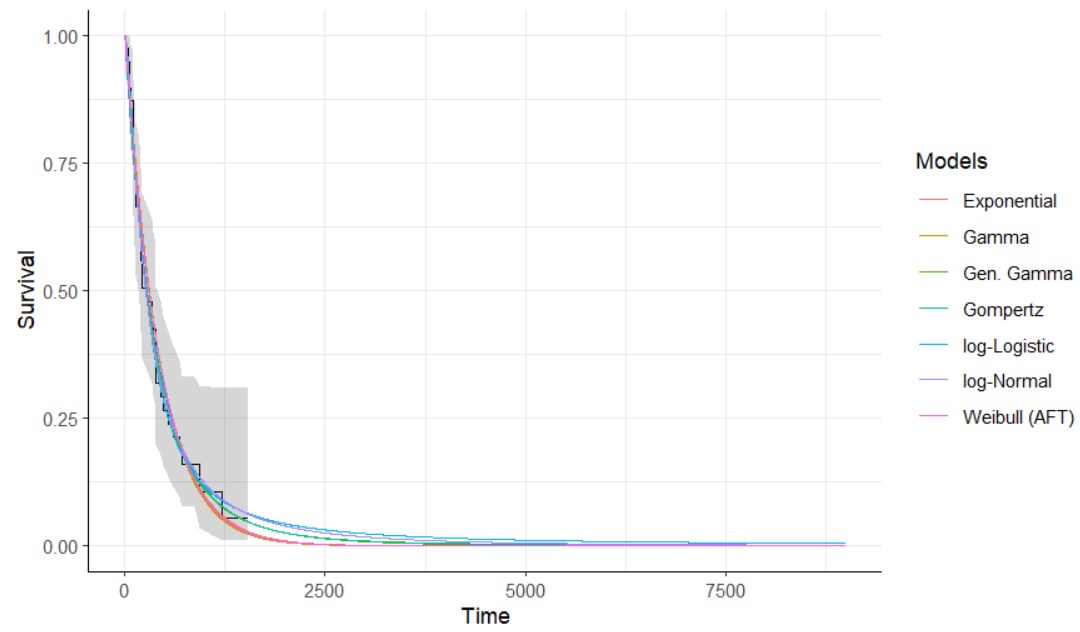

**Supplementary Figure 19:**

**ALK 2L-BSC:**

| Distribution | AIC_PFS | Rank_PFS.x | BIC_PFS | Rank_PFS.y |
| --- | --- | --- | --- | --- |
| weibull | 240.8705 | 3 | 243.3867 | 3 |
| llogis | 242.1582 | 5 | 244.6744 | 5 |
| lognormal | 244.7572 | 7 | 247.2734 | 7 |
| exp | **240.2164** | **1** | **241.4744** | **1** |
| gamma | 240.6379 | 2 | 243.1541 | 2 |
| gengamma | 242.4997 | 6 | 246.274 | 6 |
| gompertz | 241.7821 | 4 | 244.2983 | 4 |

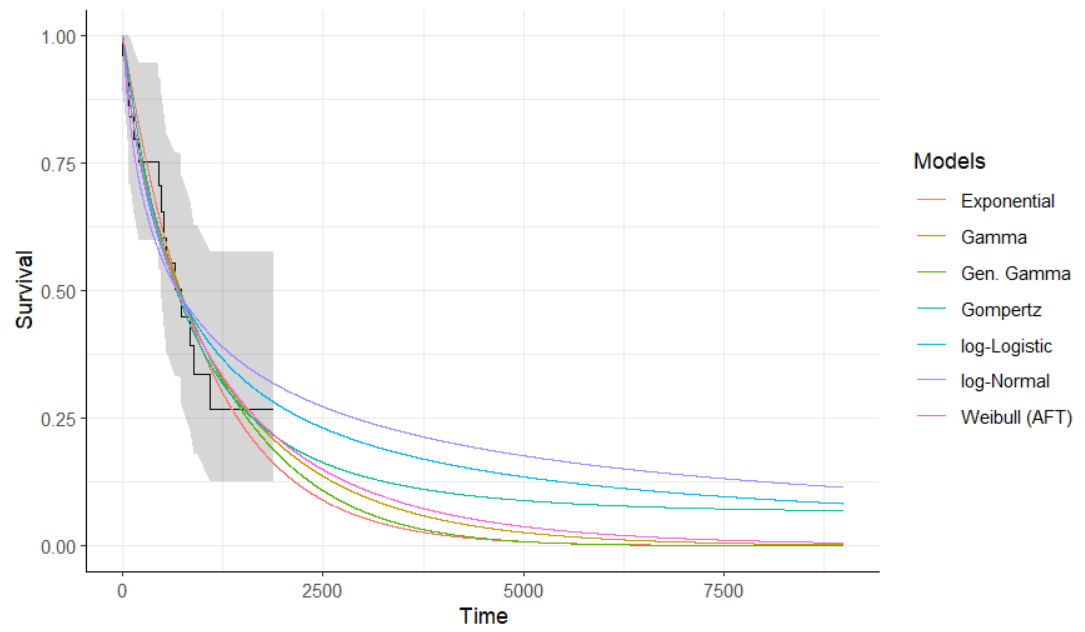

**Supplementary Figure 20:**

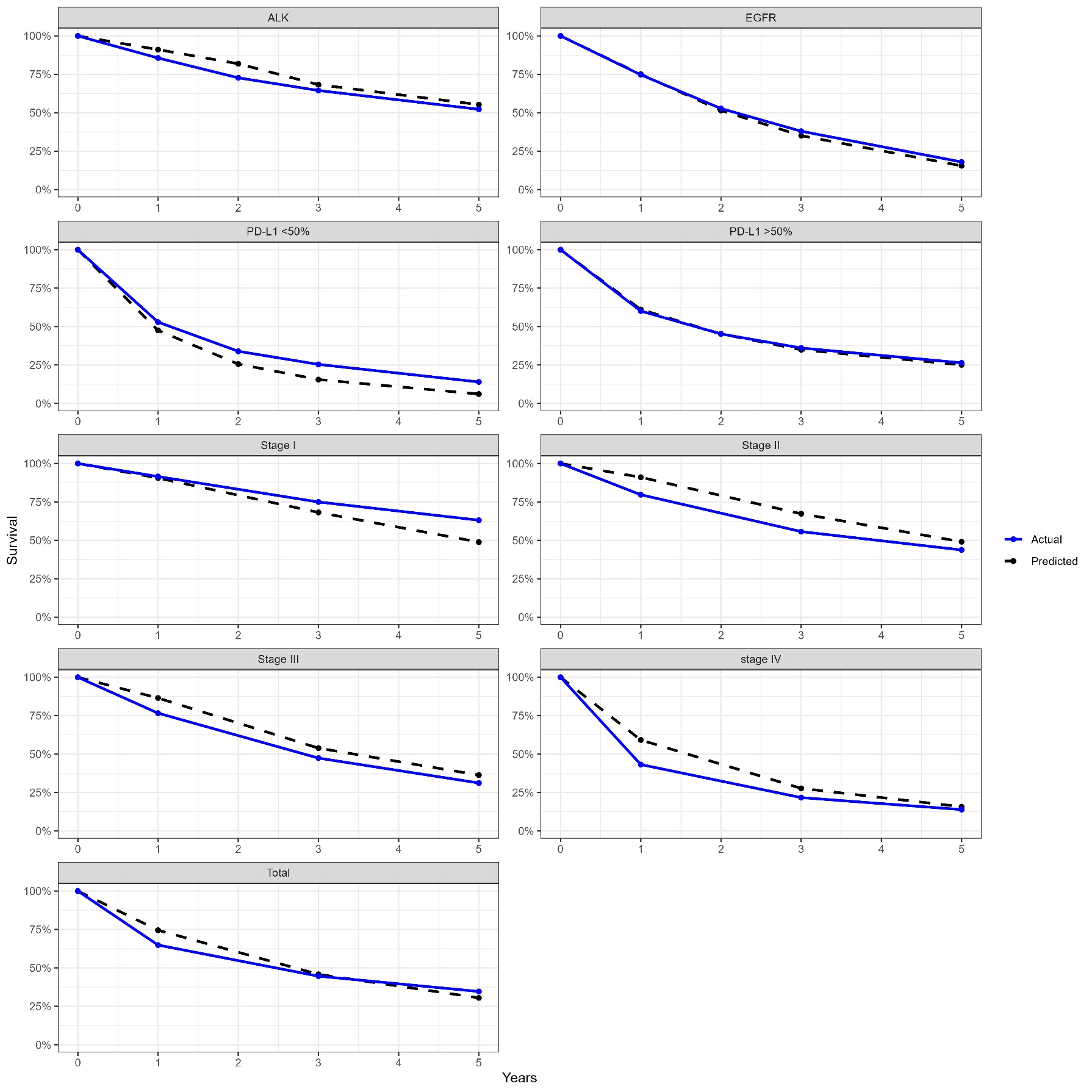

**Supplementary Figure 21:** Model-predicted (black) vs averaged observed (blue) survival probabilities in subgroups at time points

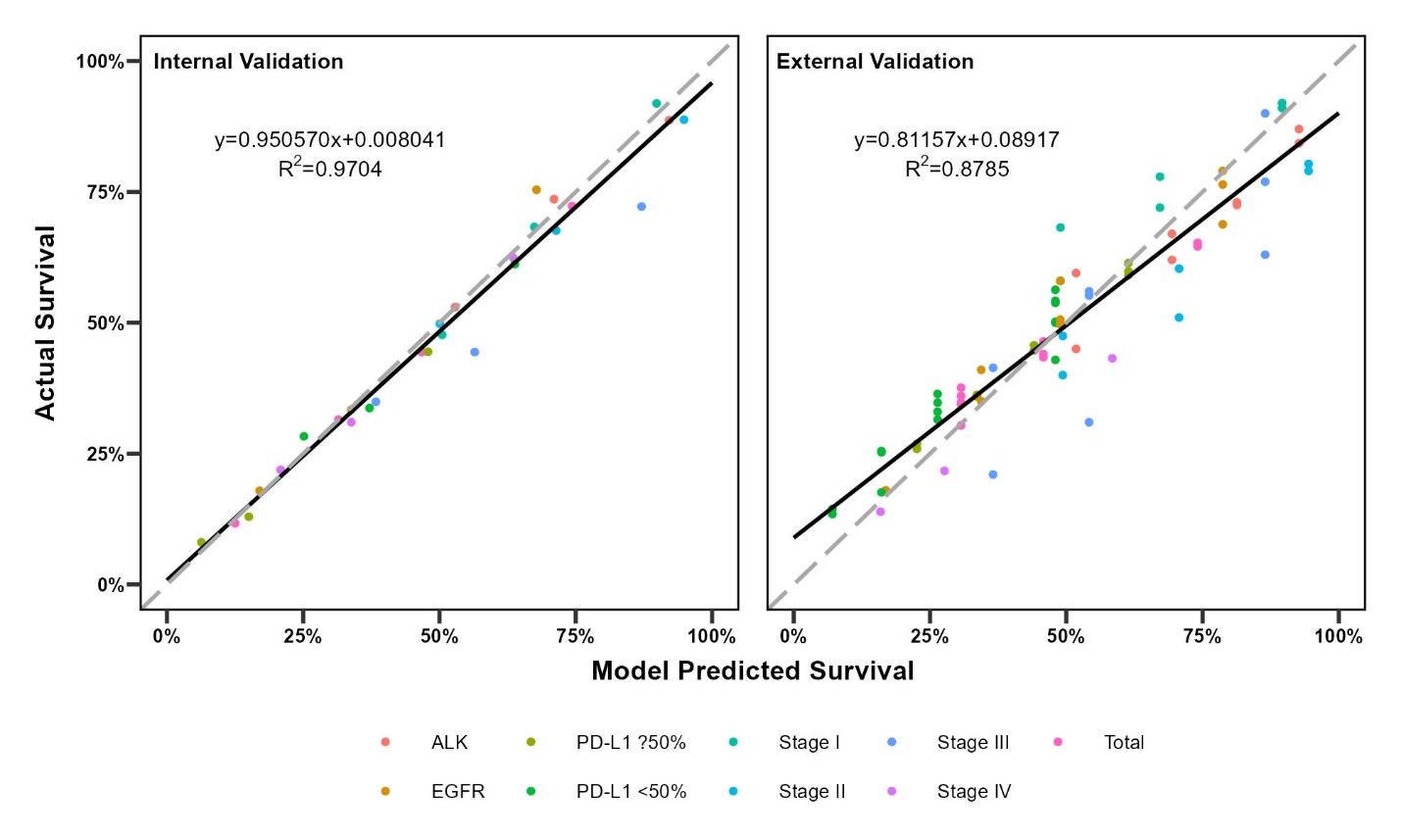

**Supplementary Figure 22:** Scatter plots and fitted linear regression results for internal validation and external validation of survival probabilities in the alternate model against the GLR cohort and observational results, where the black line represents the fitted model and the grey dashed line represents a 45-degree reference line.

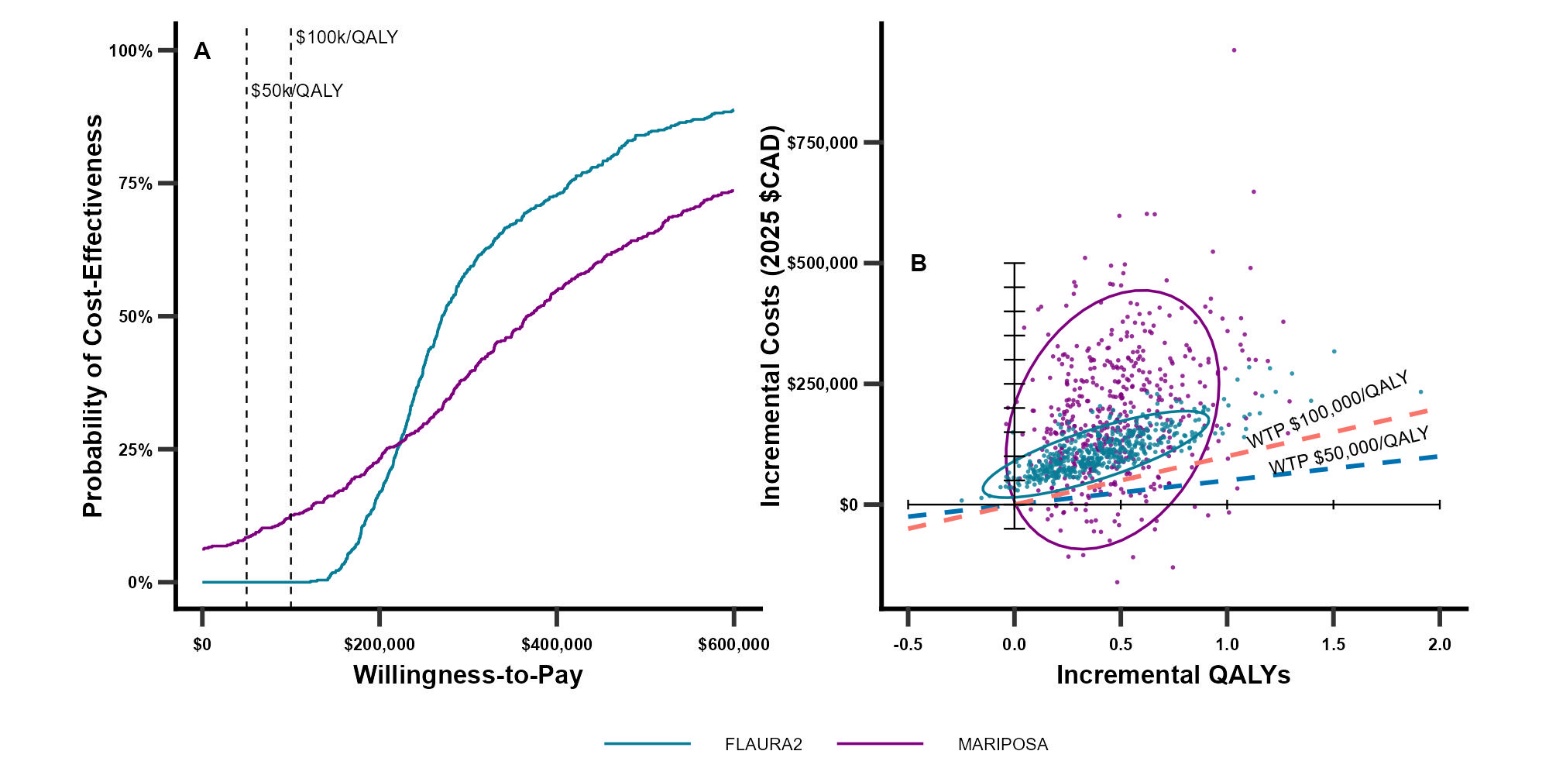

**Supplementary Figure 23:** Cost-effectiveness acceptability curves (**A**) and cost-effectiveness acceptability plane and 95% confidence ellipse (**B**) with osimertinib plus platinum pemetrexed for FLAURA2 (Blue) and amivantamab plus lazertinib for MARIPOSA (Purple) against osimertinib monotherapy for the 50% uncertainty scenario.

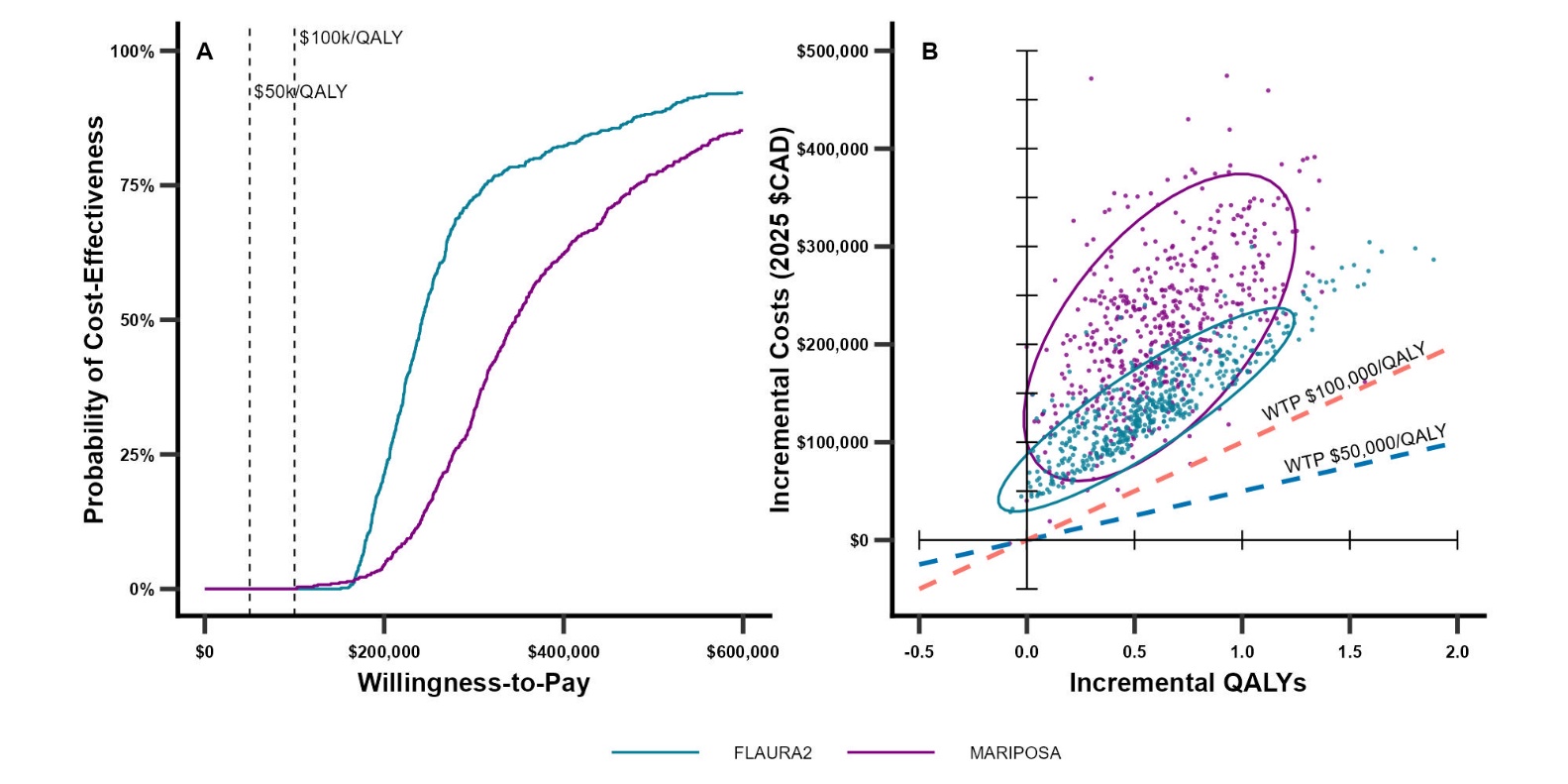

**Supplementary Figure 24:** Cost-effectiveness acceptability curves (**A**) and cost-effectiveness acceptability plane and 95% confidence ellipse (**B**) with osimertinib plus platinum pemetrexed for FLAURA2 (Blue) and amivantamab plus lazertinib for MARIPOSA (Purple) against osimertinib monotherapy in the alternate model.
